# Joint Associations of Outdoor Nitrogen Dioxide and Temperature with Incident Adult-Onset Asthma in the United States

**DOI:** 10.64898/2026.05.15.26353311

**Authors:** Shelton Lo, Gabriel A. Goodney, Hantao Wang, Jungeun Lim, Susan Valerie Czach, Jared A. Fisher, Maryam Hashemian, Rena R. Jones, Jason Y.Y. Wong

## Abstract

**Background:** Nitrogen dioxide (NO_2_) is a surrogate for traffic and industrial air pollution associated with adverse respiratory outcomes. Whether elevated NO_2_ and temperature jointly influence adult-onset asthma (AOA) risk is unclear, especially among subgroups with varying lifestyle and exposure profiles. We investigated further in the prospective All of Us research program.

**Methods:** Among 596,926 U.S. participants who consented to electronic health record release, annual average NO_2_ concentrations from satellite data were linked to residential locations for 376,535 individuals. We used multivariable Cox regression to estimate associations between NO_2_, temperature, and incident AOA, adjusting for co-pollutants and potential confounders. We analyzed 4-category cross-classification variables between NO_2_ (“high”>75^th^ percentile vs. “low” ≤75^th^ percentile) and maximum or average temperature (“high”>median vs. “low”≤median). We also stratified by sex, age, income, and smoking status. Additive interactions were estimated using Relative Excess Risk due to Interaction, Attributable Proportion, and Synergy Index.

**Results:** We identified 10,413 incident AOA cases over an average 4-year follow-up. Participants with the highest categories of NO_2_ and temperature exposure had significantly higher risk compared to those with the lowest (HR_High NO2×High Max. Temp._=1.37, 95%CI:1.26-1.49; HR_High NO2×High Average Temp._=1.49, 95%CI:1.38-1.61). The joint association of high NO_2_ and high maximum temperature was more pronounced among ever-smokers (HR=1.59, 95%CI:1.40-1.81) than never-smokers (HR=1.26, 95%CI:1.13-1.41). Interaction analyses supported super-additive interactions of high NO_2_ and high average temperature on AOA risk, particularly among ever smokers, lower-income participants, and younger adults.

**Conclusion:** Our findings highlight the respiratory health threat of long-term joint exposure to elevated NO₂ and average temperature, particularly among vulnerable subgroups.

## Introduction

Adult-onset asthma (AOA) is a chronic lower respiratory disease of significant public health concern in the United States (U.S.), as it is associated with reduced quality of life, increased healthcare costs, and increased mortality risk (1). AOA is characterized by airway inflammation and airflow obstruction that develops during adulthood and has considerably different genetic, lifestyle, and environmental risk factors compared to childhood asthma (1–3). Although overall asthma prevalence has remained relatively stable in recent years, studies have shown that AOA accounts for a large proportion of new diagnoses (2, 3). Currently, an estimated 10 million U.S. adults are diagnosed with AOA, and approximately 7% of new cases are diagnosed in adulthood each year (4, 5).

There are several key risk factors contributing to the burden of AOA in the U.S., including exposures to environmental pollutants over the life course (6–8). Tobacco smoke, air pollution, occupational exposures, and respiratory infections are important risk factors that influence disease pathogenesis, especially when comparing AOA to childhood asthma (9, 10). Notably, environmental pollutants such as fine particulate matter (PM_2.5_) and nitrogen dioxide (NO_2_) have been suggested in promoting airway inflammation and asthma exacerbation in adults, including individuals with no history of asthma (11–13).

Recent studies have identified key demographic differences in the prevalence and severity of AOA, with higher rates observed among certain racial and ethnic groups, individuals of lower socioeconomic status, and groups residing in regions with elevated environmental pollutant levels (14–16). Outdoor air pollution, PM_2.5_ in particular, has been found to contribute to elevated AOA risk and increased disease severity (12, 13). Furthermore, there is some evidence suggesting that NO_2_ can trigger asthma-like symptoms and potentially increase the risk of developing AOA. However, the joint effects of NO_2_ and temperature on AOA risk remain poorly understood (17, 18) and findings have been inconsistent across different populations and geographic features when accounting for other air pollutants and atmospheric conditions (19). Importantly, NO_2_ is both a major primary emission from traffic combustion sources and a commonly used surrogate of traffic- and industry-related air pollution mixtures. The strong correlation of NO_2_ with co-emitted pollutants from these sources may partly explain the complex patterns of association observed in AOA risk (20). NO_2_ formation is closely linked with temperature through photochemical reactions, and understanding how chronic exposure to NO_2_ may impact AOA development across diverse demographic groups is an important area of investigation (21).

To address these knowledge gaps, we leveraged data from the All of Us research program (AoU), a prospective cohort of U.S. volunteers (22). Here, we examined the associations between residential NO_2_, temperature, and risk of incident AOA. Our work aims to disentangle the complex relations between atmospheric conditions and AOA, and to identify subgroups that may be especially susceptible to the harmful respiratory effects of environmental pollutants. Importantly, most prior studies have not formally evaluated whether NO_2_ and temperature interact on the additive scale to influence AOA risk, which is arguably the most relevant interaction for joint exposure interventions (17, 18). Our study has potentially important public health implications by informing future research on targeted exposure reduction strategies to reduce the burden of chronic lower respiratory diseases in the United States, particularly in vulnerable subgroups.

## Methods

### Study Population

The AoU is a large-scale prospective cohort study that enrolled over 848,000 volunteers aged ≥18 years from across the U.S. (allofus.nih.gov) (22). In the current analyses, we included 633,547 participants who met inclusion criteria of being ≥18 years old, residing within the U.S. or its territories, providing informed consent, and who were followed prospectively between 2017 and 2023. Demographic variables such as date of birth, sex, race, Hispanic ethnicity, educational attainment, annual household income, and smoking status were collected through self-reported questionnaires.

Medical information was extracted from electronic health records (EHR) obtained from partner healthcare organizations and standardized according to the Observational Medical Outcomes Partnership (OMOP) Common Data Model. Among the eligible participants, 596,926 consented to EHR data release. Geolocation information was derived from participant-provided addresses or EHR data, with residential history gathered at enrollment through the “Basics” survey. To ensure participant privacy, residential geolocation data were limited to three-digit ZIP code regions. Furthermore, postal regions with <20,000 participants were aggregated into a nearby three-digit ZIP code. After excluding participants without EHR data, those with prevalent asthma diagnosed prior to enrollment, and individuals lacking valid residential history, the final analytic cohort consisted of 376,535 participants. Written informed consent was obtained from all participants, and data were fully anonymized before analysis. All analyses were performed using the AoU Controlled Tier Dataset v8 (C2024Q3R4) released in February 2025.

### Exposure Assessment and Data Linkage

Ambient air pollutant estimates derived from high-resolution ensemble model predictions were obtained from NASA’s Socioeconomic Data and Applications Center (SEDAC) and Earth Observing System Data and Information System (EOSDIS). These models combined satellite-derived, meteorological, and land-use data to estimate daily and annual concentrations of fine particulate matter (PM_2.5_; µg/m³), ozone (ppb), and NO_2_ (ppb) at the ZIP code level across the contiguous U.S. from 2000 to 2016 (23, 24). Annual pollutant levels were aggregated at the three-digit ZIP code level by averaging values from constituent five-digit ZIP codes.

Because temperature can influence atmospheric chemistry, pollutant dispersion, and respiratory susceptibility, we also extracted annual minimum, maximum, and average temperature data from the National Oceanic and Atmospheric Administration’s National Centers for Environmental Information (NOAA/NCEI) (25–27). These temperature metrics were spatially linked to participants’ residential ZIP codes.

Geographical coordinates for ZIP codes were obtained from the 2010 Census using the Python package uszipcode (version 1.0.15-8). To assign regional exposure levels while maintaining participant confidentiality, we utilized geographic centroids for each 3-digit ZIP code. For each centroid, we identified the five nearest-neighbor monitoring stations based on Haversine distance.

Environmental exposures were assigned using participants’ reported residential addresses at enrollment and their reported duration of residence at those addresses. For each participant, pollutant and temperature estimates were averaged over the calendar years that overlapped with both the participant’s reported residence period and the available environmental data period. Participants without exposure estimates for the relevant residence years were excluded from exposure-specific analyses.

### Spearman’s Correlation Analysis

We evaluated Spearman correlations (r_s_) among independent variables. Modest inverse correlations were observed between ozone and PM_2.5_ (r_s_ = -0.29), ozone and NO_2_ (r_s_ = -0.30), annual maximum temperature and PM_2.5_ (r_s_ = -0.16), and annual average temperature and PM_2.5_ (r_s_ = -0.08). Positive correlations were observed between PM_2.5_ and NO_2_ (r_s_ = 0.41), NO_2_ and annual average temperature (r_s_ = 0.17), ozone and annual average temperature (r_s_ = 0.48), NO_2_ and annual maximum temperature (r_s_ = 0.08), ozone and annual maximum temperature (r_s_ = 0.60), as well as annual average and maximum temperatures (r_s_ = 0.97).

### Outcome Assessment

The primary outcome was incident AOA, defined as the first recorded diagnosis of asthma occurring at age ≥18 years. This was ascertained through linked EHR data using International Classification of Diseases, Tenth Revision (ICD-10) code: J45.909 (28) and Systematized Nomenclature of Medicine-Clinical Terms (SNOMED CT) code: 233679003 (29).

### Statistical Analysis

#### Descriptive Analyses

Descriptive statistics were used to summarize participant characteristics. Means and standard deviations were reported for continuous variables, and differences among groups were assessed using ANOVA and t-tests. Frequencies and percentages were reported for categorical variables, and differences among groups were assessed using chi-squared tests. Choropleth (“heat”) maps were generated using R (ggplot2, tmap) to visualize geographic patterns of adult-onset asthma incidence across the U.S. Crude incidence rates were calculated using individual-level EHR data over the follow-up period. Subsequently, the 2000 U.S. standard population weights were used to estimate age-adjusted incidence rates. For visualization, age-adjusted incidence rates were aggregated to the state level using direct standardization and displayed on the Choropleth maps.

#### Exposure-Disease Association Analyses

Multivariable Cox proportional hazards models were used to estimate hazard ratios (HRs) and 95% confidence intervals (CIs) for incident AOA in relation to NO*_2_* exposure, modeled both continuously (per 8 ppb) and by quartiles, as well as annual maximum and average temperature, modeled continuously (per 1 °C). Time-on-study (years) was used as the time scale. We adjusted for age at enrollment (years), sex (male, female), race (White, Black or African American, Asian, Other), Hispanic ethnicity, annual household income (“low” <$75,000, “mid” $75,000–$150,000, “upper” >$150,000), educational attainment (high school graduate or less, some college, college graduate, or advanced degree or higher), BMI (kg/m²), smoking status (never- or ever-smoker), smoking pack-years (py), ozone concentrations (ppb), and PM_2.5_ concentrations (µg/m³).

To evaluate the joint associations of residential NO_2_ concentrations and temperature on the risk of AOA, NO_2_ levels were dichotomized at the highest quartile to define low and high exposure groups (≤75^th^ percentile vs. >75^th^ percentile ppb), and similarly, annual average and maximum temperatures were categorized into low and high groups (≤ median vs. > median °C). These dichotomized variables were then cross-classified to create a four-level categorical variable representing joint exposure:

low NO_2_/low temperature (reference category), high NO_2_/low temperature, low NO₂/high temperature, and high NO_2_/high temperature. Additionally, we conducted stratified analyses to evaluate heterogeneity among demographic subgroups defined by age, sex, smoking status, and annual household income as a proxy for socioeconomic status. The subgroup analyses were specified *a priori*. Subsequently, we fit multiplicative interaction terms between NO_2_ x temperature, age, sex, smoking status, and annual household income in the Cox models to test for heterogeneity. Proportional hazards assumptions were visually assessed via Schoenfeld residuals, with no violations detected for the main effect of NO_2_ and temperature. Two-sided P-values <0.05 were considered statistically significant.

#### Additive Interaction Analyses

We estimated three measures of additive interaction from the four-level NO_2_ × temperature cross-classification variables: the Relative Excess Risk due to Interaction (RERI), the Attributable Proportion (AP), and the Synergy Index (S), with 95% CIs estimated using 100,000-iteration Monte Carlo simulation (30). RERI estimates how much the joint HR exceeds what would be expected from adding the two individual exposure HRs. AP rescales RERI as a fraction of the joint risk, expressing the proportion of excess risk in the high NO_2_ - high temperature group that is due to the interaction. Synergy Index (S) compares the joint excess risk to the sum of individual excess risks. RERI > 0, AP > 0, and S > 1 indicate super-additive interaction, where the joint association of high NO_2_ and high temperature on AOA risk exceeds the sum of their independent effects (30).

#### Accounting for Potential Volunteer Selection Bias

The AoU research program is not a population-based study with probabilistic sampling. Rather, AoU enrolled volunteers, which helps maximize the potential to recruit a large number of participants needed to investigate complex interactions in the omics era (31–33). To account for potential volunteer bias in participant enrollment, cell weighting was applied to each participant in the statistical models by calibrating the marginal distribution of various demographic factors using the 2018-2022 American Community Survey (ACS) Public Use Microdata Sample (PUMS) as a reference. The weight flooring and ceiling were set at the 5^th^ and 95^th^ percentiles. The first set of weights adjusted for age, race, and Hispanic ethnicity at the state level. The second set of weights contained the variables from the first set and additionally adjusted for education and income level.

#### Sensitivity Analyses - Quantile-Adaptive Sure Independence Screening (QaSIS) Method

To rank the influence of 18 predictors (i.e., air pollutants, temperature, and each covariate) on quantiles of time-to-diagnosis for AOA, we applied the quantile-adaptive sure independence screening (QaSIS) method for censored data on complete-case participants (34). Because AOA is a rare event in our sample, we screened at a high-survival probability (τ=0.98), which corresponds to an early time-to-diagnosis. Detailed results are presented in Supplementary Tables.

#### Sensitivity Analyses – Censored Quantile Regression Analysis

To assess the influence of NO_2_ and temperature on extreme time-to-diagnosis, we conducted a censored quantile regression (CQR) sensitivity analysis on complete-case participants (35–37). Because AOA was rare in our sample, only the earliest survival quantiles were identifiable; therefore, CQR was restricted to tau=0.02, corresponding to the earliest 2% of diagnosis times among incident AOA cases. These results were compared to HR estimates from Cox models to assess whether the direction of the associations were consistent.

## Results

### Baseline Characteristics

The distribution of key baseline characteristics is shown in Table 1. At enrollment, those who developed incident AOA cases during the follow-up were younger (49.6 years vs. 52.1 years), had a larger proportion of female participants (68.8% vs. 60.2%), and had a larger proportion of Black participants (23.0% vs. 18.0%) compared to non-cases throughout the follow-up. Additionally, those who developed incident AOA cases during the follow-up had lower educational attainment (32.6% vs. 26.9% with high school education or less) and household incomes (48.0% vs. 38.1% earning <$75,000 per year). Tobacco use patterns differed slightly between both groups -never-smokers made up 54.7% of incident AOA cases compared to 57.9% among non-cases. AOA cases had a higher proportion of ever-smokers (42.7% vs. 38.7%) and reported higher average pack-years (py) of smoking (6.3 py) compared to non-cases (5.4 py).

**Table 1:** Characteristics of Study Participants from the All of Us Research Program at Enrollment. (**2017–2023**)

| Characteristic | Overall Study Sample | Incident adult-onset asthma case diagnosed during follow-up | Non-case throughout the follow-up |
| --- | --- | --- | --- |
| Number of participants | 376535 | 10413 | 366122 |
| Age (Years) (mean, SD) | 52.0 (17.0) | 49.6 (16.5) | 52.1 (17.0) |
| Age at diagnosis of adult-onset asthma (mean, SD) | - | 51.2 (16.6) | - |
| Sex |  |  |  |
| Male (n, %) | 142377 (37.8) | 3048 (29.3%) | 139329 (38.1) |
| Female (n, %) | 227454 (60.4) | 7167 (68.8) | 220287 (60.2) |
| Race |  |  |  |
| White (n, %) | 216153 (57.4) | 5455 (52.4) | 210698 (57.5) |
| Black (n, %) | 68478 (18.2) | 2394 (23.0) | 66084 (18.0) |
| Asian (n, %) | 11620 (3.1) | 230 (2.2) | 11390 (3.1) |
| Ethnicity |  |  |  |
| Hispanic or Latino (n, %) | 68351 (18.2) | 2030 (19.5) | 66321 (18.1) |
| Not Hispanic or Latino (n, %) | 297159 (78.9) | 8066 (77.5) | 289093 (79.0) |
| Smoking Status |  |  |  |
| Ever smoker (n, %) | 146107 (38.8) | 4445 (42.7) | 141662 (38.7) |
| Never smoker (n, %) | 217652 (57.8) | 5693 (54.7) | 211959 (57.9) |
| Smoking pack-years (py) (mean, SD)* | 5.4 (13.9) | 6.3 (14.9) | 5.4 (13.9) |
| Body mass index (kg/m<sup>2</sup>) (mean, SD) | 29.9 (7.6) | 31.7 (8.5) | 29.8 (7.6) |
| Educational Attainment |  |  |  |
| High school, GED, or below | 102048 (27.1) | 3393 (32.6) | 98655 (26.9) |
| Undergraduate | 97198 (25.8) | 2951 (28.3) | 94247 (25.7) |
| College | 86645 (23.0) | 2105 (20.2) | 84540 (23.1) |
| Advanced degree | 81516 (21.6) | 1716 (16.5) | 79800 (21.8) |
| Annual household income |  |  |  |
| Below \$75,000 | 144593 (38.4) | 4994 (48.0) | 139599 (38.1) |
| \$75,000 to \$150,000 | 112752 (29.9) | 2519 (24.2) | 110233 (30.1) |
| Above \$150,000 | 44440 (11.8) | 838 (8.0) | 43602 (11.9) |
| Duration of Residence at Baseline, n (%) |  |  |  |
| 0 to 2 years | 121327 (32.3) | 4106 (39.4) | 117221 (32.0) |
| 3 to 5 years | 66864 (17.8) | 1823 (17.5) | 65041 (17.8) |
| 6 to 10 years | 51827 (13.8) | 1316 (12.6) | 50511 (13.8) |
| 11 to 20 years | 57179 (15.2) | 1411 (13.6) | 55768 (15.2) |
| >20 years | 73879 (19.6) | 1623 (15.6) | 72256 (19.7) |
| Annual average temperature<br>(°C) (Mean, SD) | 14.4 (5.2) | 15.0 (5.2) | 14.4 (5.2) |
| Annual fine particulate matter<br>(PM<sub>2.5</sub>) (ug/m<sup>3</sup>) (Mean, SD) | 10.6 (2.4) | 10.5 (2.5) | 10.6 (2.4) |
| Annual nitrogen dioxide (NO<sub>2</sub>)<br>(ppb) (Mean, SD) | 25.8 (9.8) | 27.2 (10.2) | 25.8 (9.7) |
| Annual ground-level ozone<br>(ppbv) (Mean, SD) | 38.7 (4.3) | 38.8 (4.6) | 38.7 (4.3) |

\*Based on an overall sample size of 321,025 participants with smoking data.
Discrepancy in counts and proportions was due to missing/unknown data.
The mean (SD) age at AOA diagnosis is 46.4 (16.7) years when combining prevalent and incident AOA cases.

Residential duration at baseline did not differ greatly between incident AOA cases and the overall study population **(Table 1)**. AOA cases were exposed to similar annual ambient ozone concentrations at their residential location (38.8 ppb) compared to the overall population (38.7 ppb). However, annual average temperatures were slightly higher among AOA cases compared to the overall population (15.0°C vs. 14.4°C). Annual average NO₂ exposure was slightly higher among incident AOA cases (27.2 ppb) relative to the full cohort (25.8 ppb). Annual PM_2.5_ concentrations were similar by AOA status. NO₂ concentrations were higher in the northeast regions as well as certain areas in the southwest regions **(Figure 1A)**. This approximately corresponded with the higher age-adjusted AOA incidence rates in these states **(Figure 1B)**.

**Figure 1:**
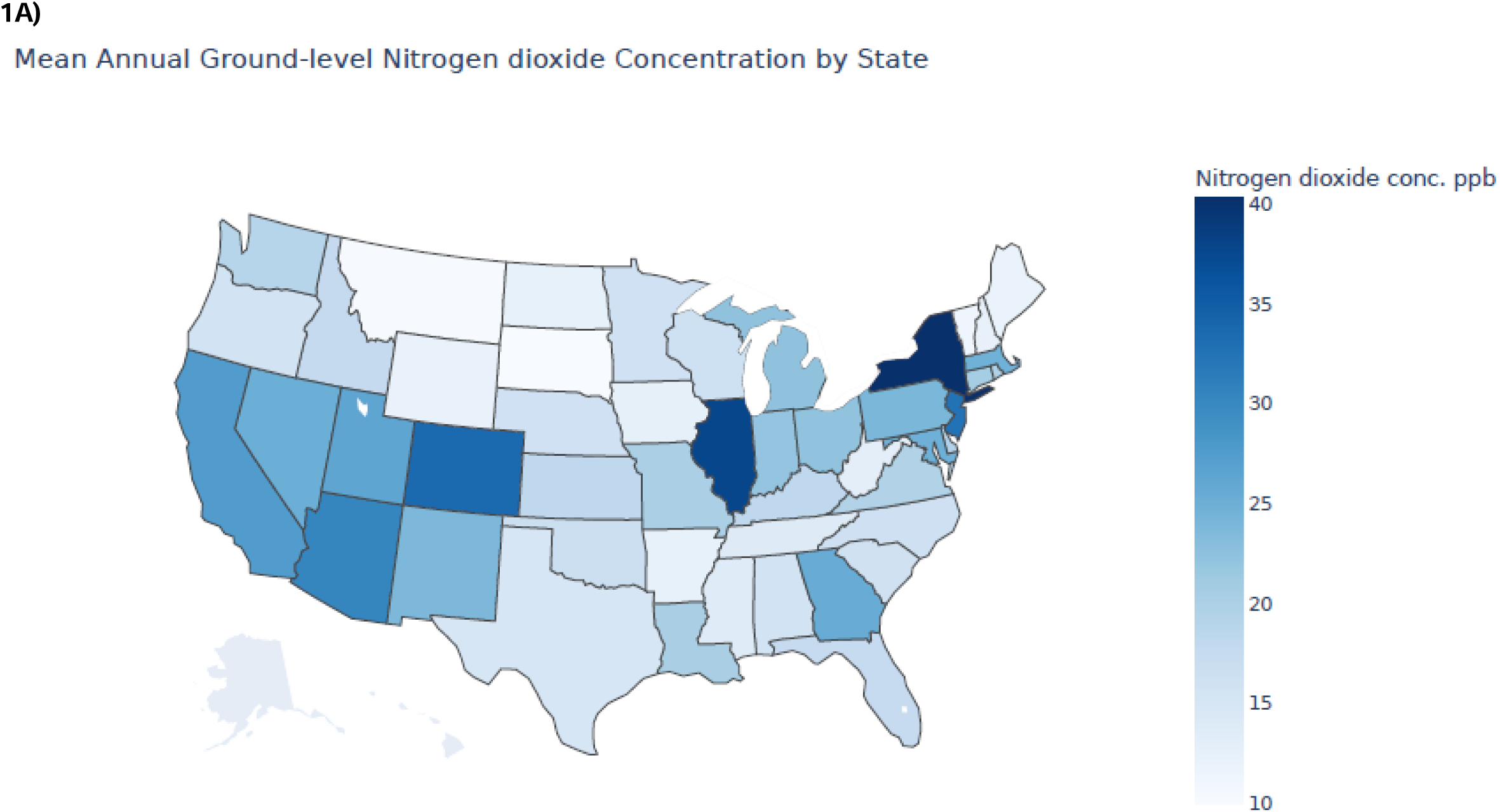
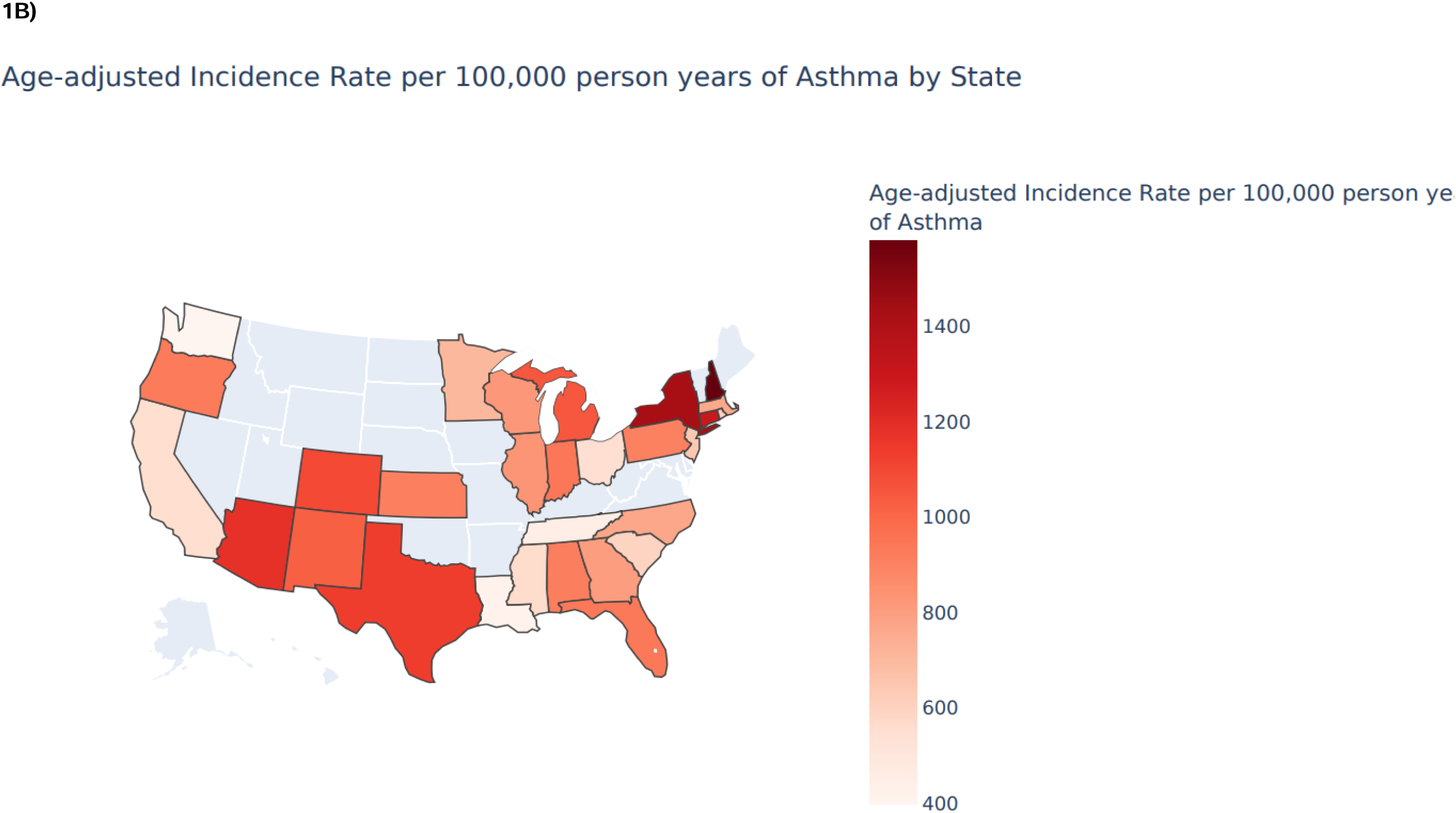
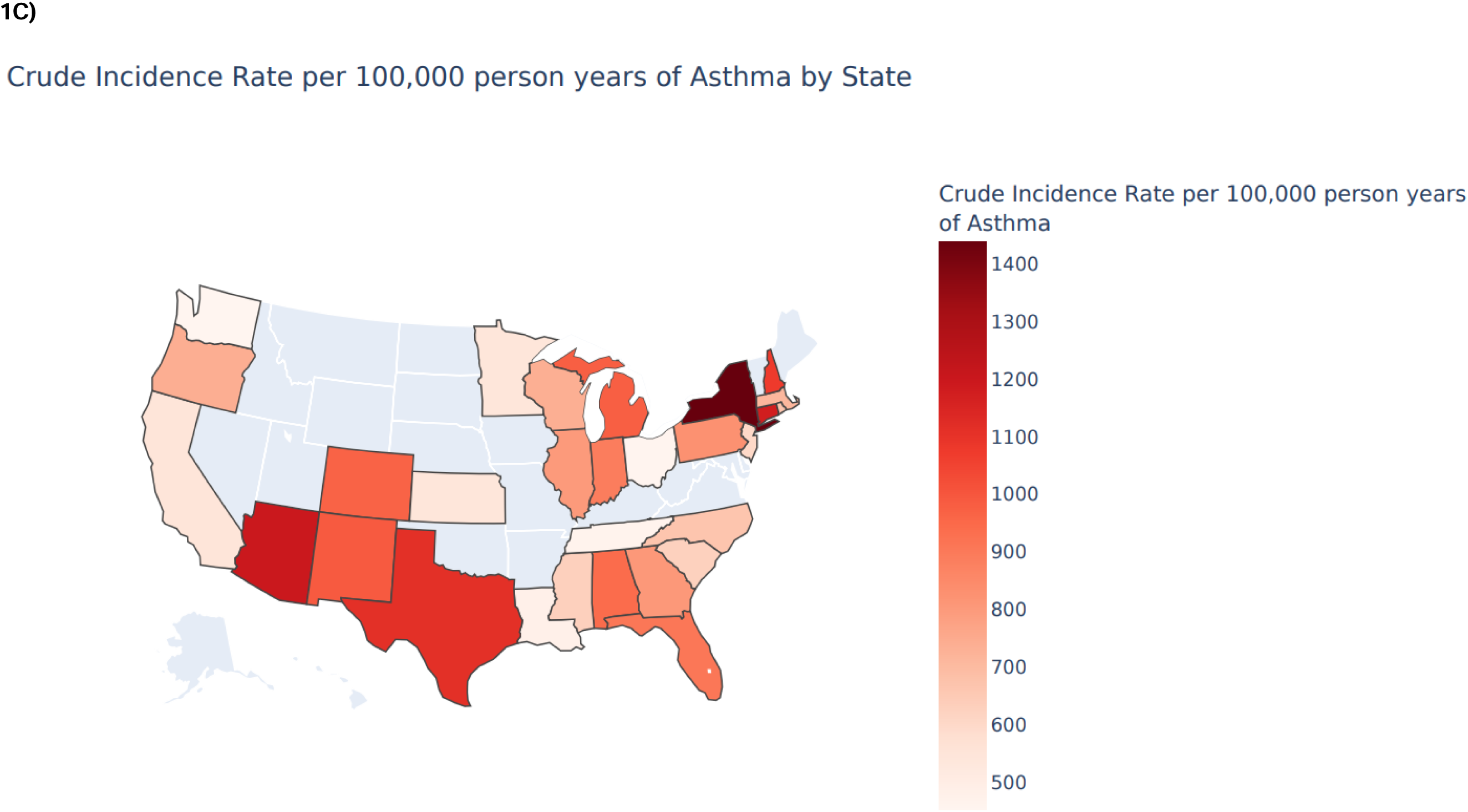
Distribution of A) NO2 concentration between 2000—2016, B) age-adjusted adult-onset asthma incidence rate, and C) crude adult-onset asthma incidence rate by state in the contiguous United States

### Overall Association between Residential NO_2_ Exposure and Risk of AOA

We identified 10,413 incident AOA cases over an average 4-year follow-up. The mean age at diagnosis was 51.2 years. We observed a positive association between annual average residential NO_2_ and AOA risk **(Supplementary Table 1)**; however, the trend was non-monotonic. This relationship was apparent only among individuals in the highest NO_2_ exposure quartile (HR: 1.08, 95%CI:1.02-1.15).

### Overall Association between Temperature and Risk of AOA

We found a modest association between exposure to higher temperatures and increased risk of AOA. Here, each 1°C increase in residential annual maximum temperature and average temperature was both associated with a 1% increase in AOA risk (HR_Maximum_= 1.01, 95%CI: 1.00-1.02; HR_Average_= 1.01, 95%CI:1.00-1.01, respectively) **(Supplementary Table 2)**. However, the trend was significant only for maximum temperature (P-trend=0.035).

### NO_2_ and AOA Association Accounting for Temperature

Our cross-variable analyses showed evidence for joint associations between high NO_2_ and temperature on AOA risk **(Table 2 and Table 3)**. Compared to individuals residing in areas with both low NO_2_ and low maximum temperature exposures (reference level), those living in high NO_2_ and high maximum temperature areas had elevated AOA risk (HR = 1.37; 95%CI:1.26-1.49; **Table 2**). Similar increased risks were observed for both low NO_2_ and high maximum temperature exposures (HR = 1.13; 95% CI: 1.06, 1.21) and high NO_2_ and low maximum temperature exposures (HR = 1.28; 95% CI: 1.17, 1.40; **Table 2**). Similarly, a significant increased risk of AOA was observed among participants exposed to high NO_2_ and high average temperatures (HR = 1.49; 95%CI:1.38-1.61) compared to participants exposed to low NO_2_ and low average temperatures **(Table 3)**. Importantly, our formal interaction analyses supported a super-additive interaction only between high NO_2_ and high average temperatures (RERI (95% CI)= 0.37(0.19,0.54); AP (95% CI)= 0.25 (0.13,0.35) (**Table 3**), but not for high NO_2_ and high maximum temperatures (**Table 2**).

**Table 2:** Nitrogen Dioxide, maximum temperature, and adult-onset asthma risk in the All of Us Research Program.

| **Ground-level Nitrogen Dioxide (NO<sub>2</sub>) and Maximum Temperature Levels** | **Cases** | **Non-Cases** | **HR (95%CI)<sup>1, 2</sup>** |
| --- | --- | --- | --- |
| Low Levels of NO<sub>2</sub> and Low Levels of Annual Maximum Temperature | 1918 | 68877 | 1.00 (Ref.) |
| Low Levels of NO<sub>2</sub> and High Levels of Annual Maximum Temperature | 1928 | 65405 | 1.13 (1.06, 1.21) |
| High Levels of NO<sub>2</sub> and Low Levels of Annual Maximum Temperature | 729 | 18555 | 1.28 (1.17, 1.40) |
| High Levels of NO<sub>2</sub> and High Levels of Annual Maximum Temperature | 912 | 23018 | 1.37 (1.26, 1.49) |

<sup>1</sup>Cox proportional hazards models were adjusted for age, sex, race, ethnicity, BMI, smoking status, smoking pack years, annual household income, educational attainment, annual O<sub>3</sub> levels, and annual PM<sub>2.5</sub> levels
<sup>2</sup>Analyses were restricted to participants with non-missing environmental and covariate data; therefore, case counts differ from the overall analytic sample.
Interaction analyses: RERI (95% CI)= -0.04(-0.22,0.14); AP (95% CI)= -0.03(-0.17,0.10); S (95% CI)= 0.90(0.57,1.46)
Abbreviations: HR=Hazard Ratio; CI=Confidence Interval; Ref=Reference Group; RERI=Relative Excess Risk due to Interaction; AP= Attributable Proportion, S=Synergy Index

**Table 3:** Nitrogen Dioxide, average temperature, and adult-onset asthma risk in the All of Us Research Program.

| **Ground-level Nitrogen Dioxide (NO<sub>2</sub>) and Average Temperature Levels** | **Cases** | **Non-Cases** | **HR (95%CI)<sup>1, 2</sup>** |
| --- | --- | --- | --- |
| Low Levels of NO<sub>2</sub> and Low Levels of Annual Average Temperature | 2012 | 72743 | 1.00 (Ref.) |
| Low Levels of NO<sub>2</sub> and High Levels of Annual Average Temperature | 1834 | 61539 | 1.14 (1.07, 1.22) |
| High Levels of NO<sub>2</sub> and Low Levels of Annual Average Temperature | 397 | 14017 | 0.98 (0.88, 1.10) |
| High Levels of NO<sub>2</sub> and High Levels of Annual Average Temperature | 1244 | 27556 | 1.49 (1.38, 1.61) |

<sup>1</sup>Cox proportional hazards models were adjusted for age, sex, race, ethnicity, BMI, smoking status, smoking pack years, annual household income, educational attainment, annual O<sub>3</sub> levels, and annual PM<sub>2.5</sub> levels
<sup>2</sup>Analyses were restricted to participants with non-missing environmental and covariate data; therefore, case counts differ from the overall analytic sample.
Interaction analyses: RERI (95% CI)= 0.37 (0.19,0.54); AP (95% CI)=0.25 (0.13,0.35); S (95% CI)= Not defined because a single HR was ≤1.
Abbreviations: HR=Hazard Ratio; CI=Confidence Interval; Ref=Reference Group; RERI= Relative Excess Risk due to Interaction; AP= Attributable Proportion, S=Synergy Index

### Joint Associations of NO_2_ × Temperature and AOA Risk by Demographic Subgroups Smoking Status

We found notable differences in the joint association between NO_2_ × temperature and AOA risk by smoking status (**Table 4**). The joint association of high NO_2_ × high maximum temperature was more pronounced among ever-smokers (HR = 1.59, 95%CI:1.40-1.81) than never-smokers (HR = 1.26, 95%CI:1.13-1.41). A similarly striking difference was observed when comparing the joint associations of high NO_2_ × high average temperature among ever-smokers (HR = 1.67, 95%CI:1.49-1.86) and never-smokers (HR =1.40, 95%CI:1.27-1.55) **(Table 4)**. Importantly, our formal interaction analyses showed consistent super-additive interaction among ever-smokers [NO_2_ x Max Temp - Ever smokers: RERI (95% CI)=0.26 (−0.03,0.55); AP (95% CI)= 0.16 (−0.02,0.32); S (95% CI)= 1.79 (0.93,4.16), NO_2_ x Avg Temp - Ever smokers: RERI (95% CI)=0.67 (0.40,0.94); AP (95% CI)=0.40 (0.26,0.52); S (95% CI)= Not defined], but not in never-smokers (**Table 4**).

**Table 4:**
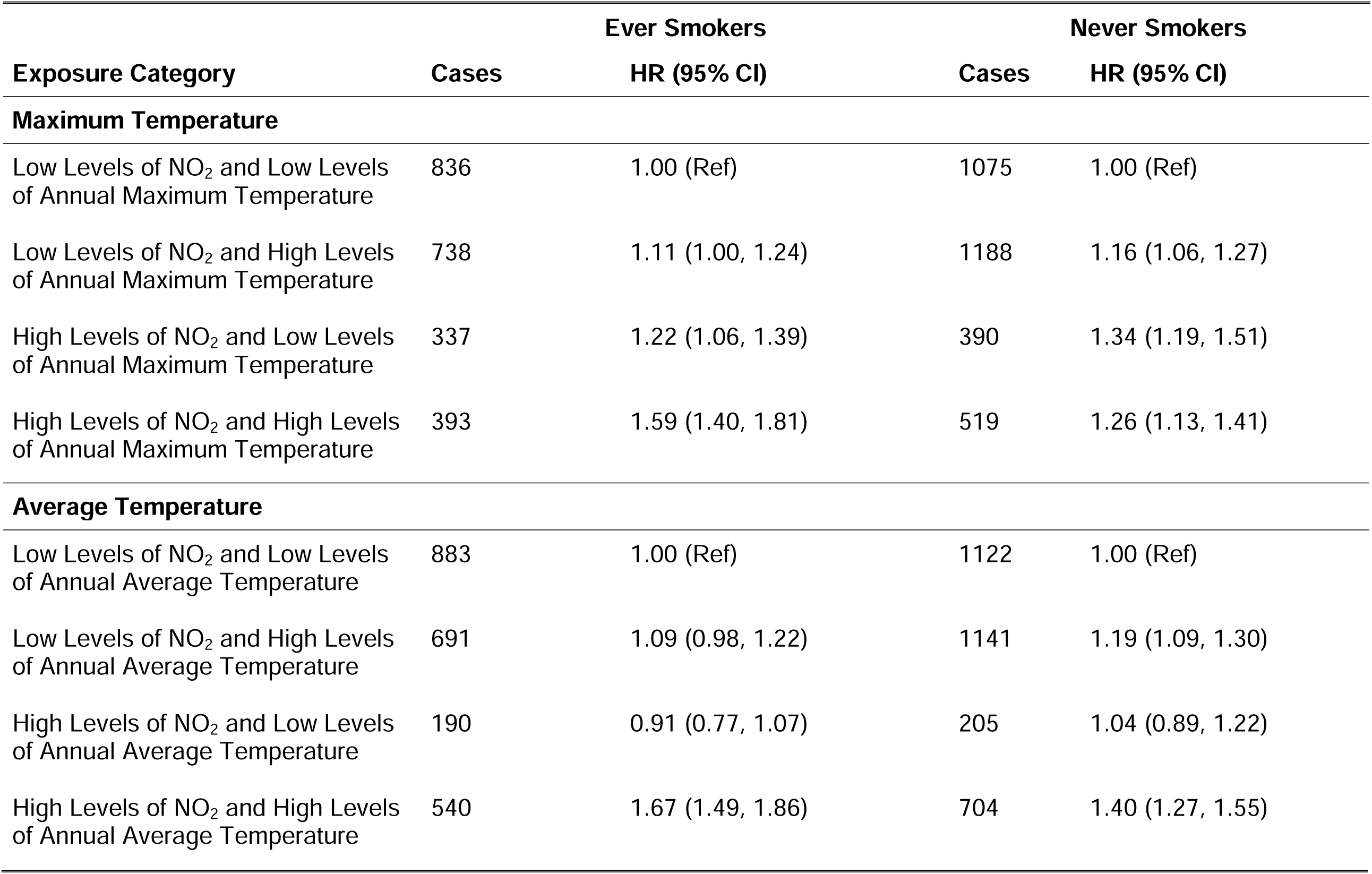
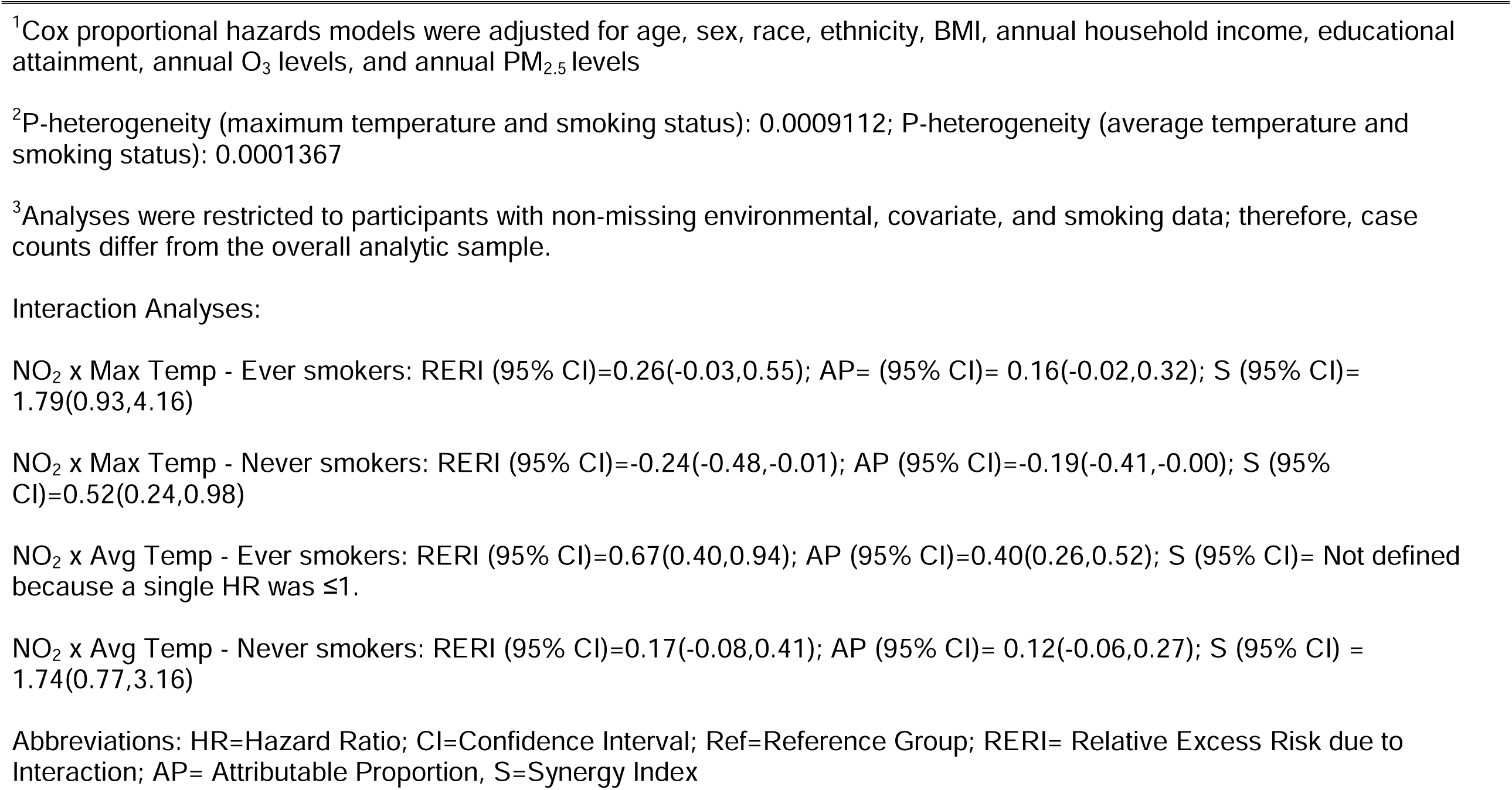
Association of nitrogen dioxide and temperature with adult-onset asthma among ever smokers and never smokers in the All of Us Research Program^1, 2, 3^.

### Sex

We found differences in the joint association between NO_2_ × temperature and AOA risk by sex **(Table 5)**. The joint association of high NO_2_ × high maximum temperature on AOA risk was higher in males (HR = 1.50, 95%CI:1.29-1.75) than females (HR = 1.33, 95%CI:1.20-1.47; P-heterogeneity=4.3x10^-14^). A similar pattern was observed with high NO_2_ × high average temperature (HR_males_=1.54, 95%CI:1.34-1.77 vs. HR_females_=1.47, 95%CI:1.36-1.61; P-heterogeneity=1.8x10^-21^;**Table 5)**. Notably, we found that the super-additive interactions between high NO_2_ and high average temperature were of greater magnitude than between high NO_2_ × high maximum temperature in both males and females (NO_2_ x Avg Temp – Males: RERI (95% CI)= 0.44 (0.11,0.76); AP=0.29 (0.08,0.45), NO_2_ x Avg Temp - Females RERI (95% CI)=0.36 (0.15,0.56); AP= 0.24 (0.11,0.36); **Table 5**).

**Table 5:**
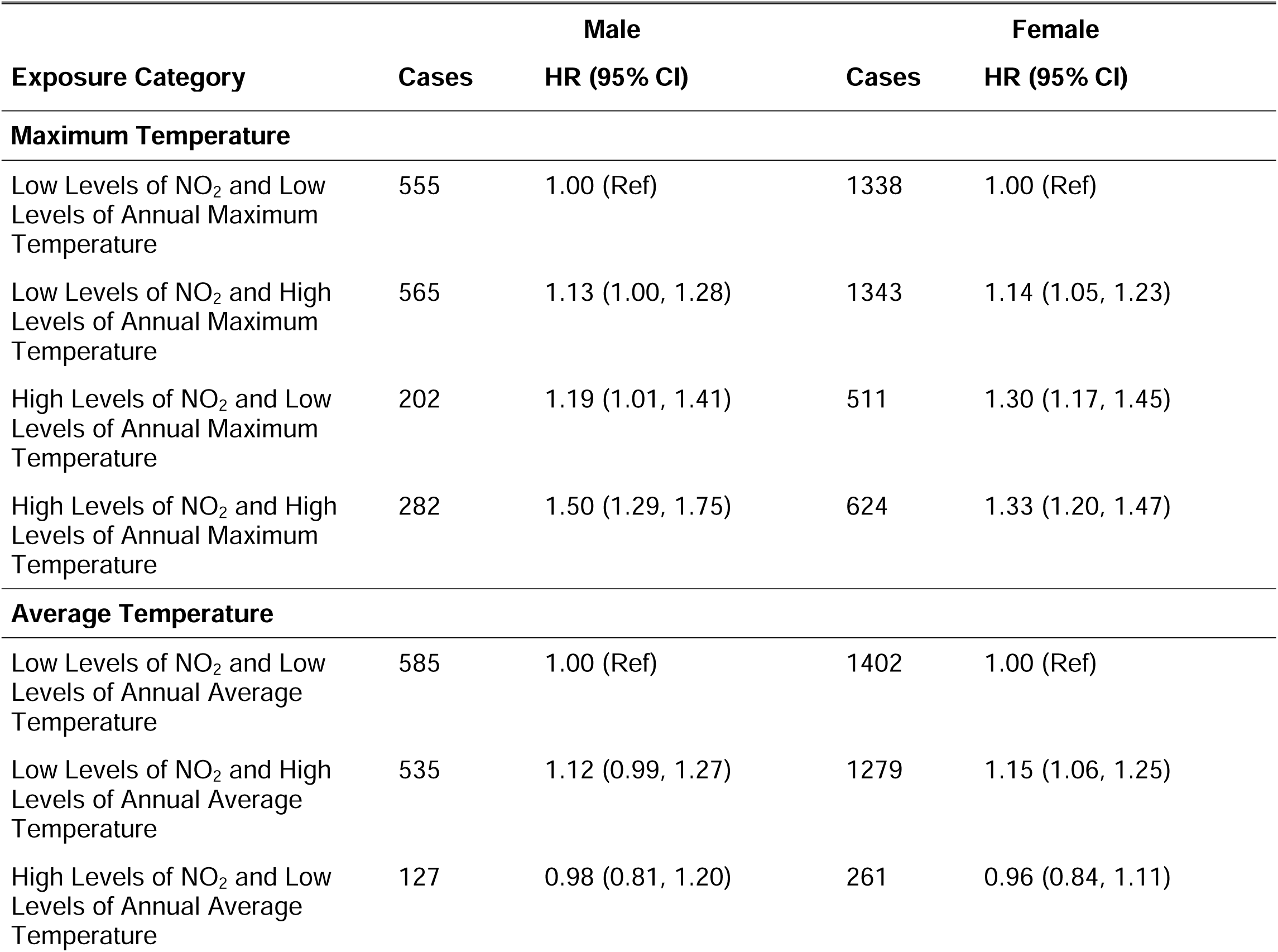
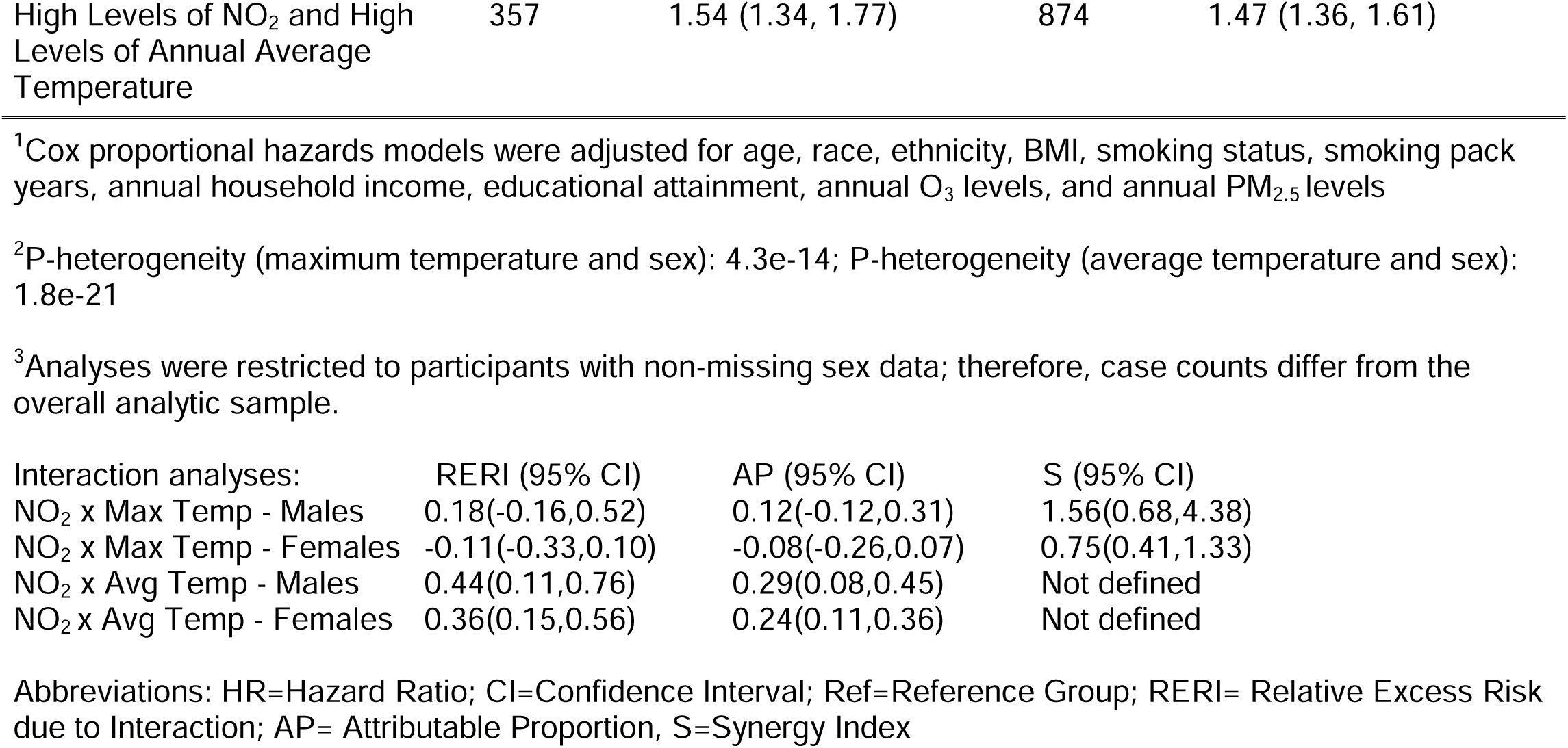
Association of nitrogen dioxide and temperature with adult-onset asthma among male and female participants in the All of Us Research Program^1, 2, 3^.

| Exposure Category | Cases | Male | Cases | Female |
| --- | --- | --- | --- | --- |
|  |  | HR (95% CI) |  | HR (95% CI) |
| Maximum Temperature |  |  |  |  |
| Low Levels of NO<sub>2</sub> and Low Levels of Annual Maximum Temperature | 555 | 1.00 (Ref) | 1338 | 1.00 (Ref) |
| Low Levels of NO<sub>2</sub> and High Levels of Annual Maximum Temperature | 565 | 1.13 (1.00, 1.28) | 1343 | 1.14 (1.05, 1.23) |
| High Levels of NO<sub>2</sub> and Low Levels of Annual Maximum Temperature | 202 | 1.19 (1.01, 1.41) | 511 | 1.30 (1.17, 1.45) |
| High Levels of NO<sub>2</sub> and High Levels of Annual Maximum Temperature | 282 | 1.50 (1.29, 1.75) | 624 | 1.33 (1.20, 1.47) |
| Average Temperature |  |  |  |  |
| Low Levels of NO<sub>2</sub> and Low Levels of Annual Average Temperature | 585 | 1.00 (Ref) | 1402 | 1.00 (Ref) |
| Low Levels of NO<sub>2</sub> and High Levels of Annual Average Temperature | 535 | 1.12 (0.99, 1.27) | 1279 | 1.15 (1.06, 1.25) |
| High Levels of NO<sub>2</sub> and Low Levels of Annual Average Temperature | 127 | 0.98 (0.81, 1.20) | 261 | 0.96 (0.84, 1.11) |
| High Levels of NO<sub>2</sub> and High Levels of Annual Average Temperature | 357 | 1.54 (1.34, 1.77) | 874 | 1.47 (1.36, 1.61) |

|  |  |  |  |
| --- | --- | --- | --- |
| Interaction analyses: | RERI (95% CI) | AP (95% CI) | S (95% CI) |
| NO<sub>2</sub> x Max Temp - Males | 0.18(-0.16,0.52) | 0.12(-0.12,0.31) | 1.56(0.68,4.38) |
| NO<sub>2</sub> x Max Temp - Females | -0.11(-0.33,0.10) | -0.08(-0.26,0.07) | 0.75(0.41,1.33) |
| NO<sub>2</sub> x Avg Temp - Males | 0.44(0.11,0.76) | 0.29(0.08,0.45) | Not defined |
| NO<sub>2</sub> x Avg Temp - Females | 0.36(0.15,0.56) | 0.24(0.11,0.36) | Not defined |

Abbreviations: HR=Hazard Ratio; CI=Confidence Interval; Ref=Reference Group; RERI= Relative Excess Risk due to Interaction; AP= Attributable Proportion, S=Synergy Index

### Age

We found that the magnitude of the joint association of high NO_2_ × high maximum temperature and AOA risk increased with age **(Table 6)**. A similar but less pronounced trend was observed for high NO_2_ × high average temperature. However, the super-additive interaction among age groups ≤65 years was more apparent for high NO_2_ × high average temperature (**Table 6**).

**Table 6:**
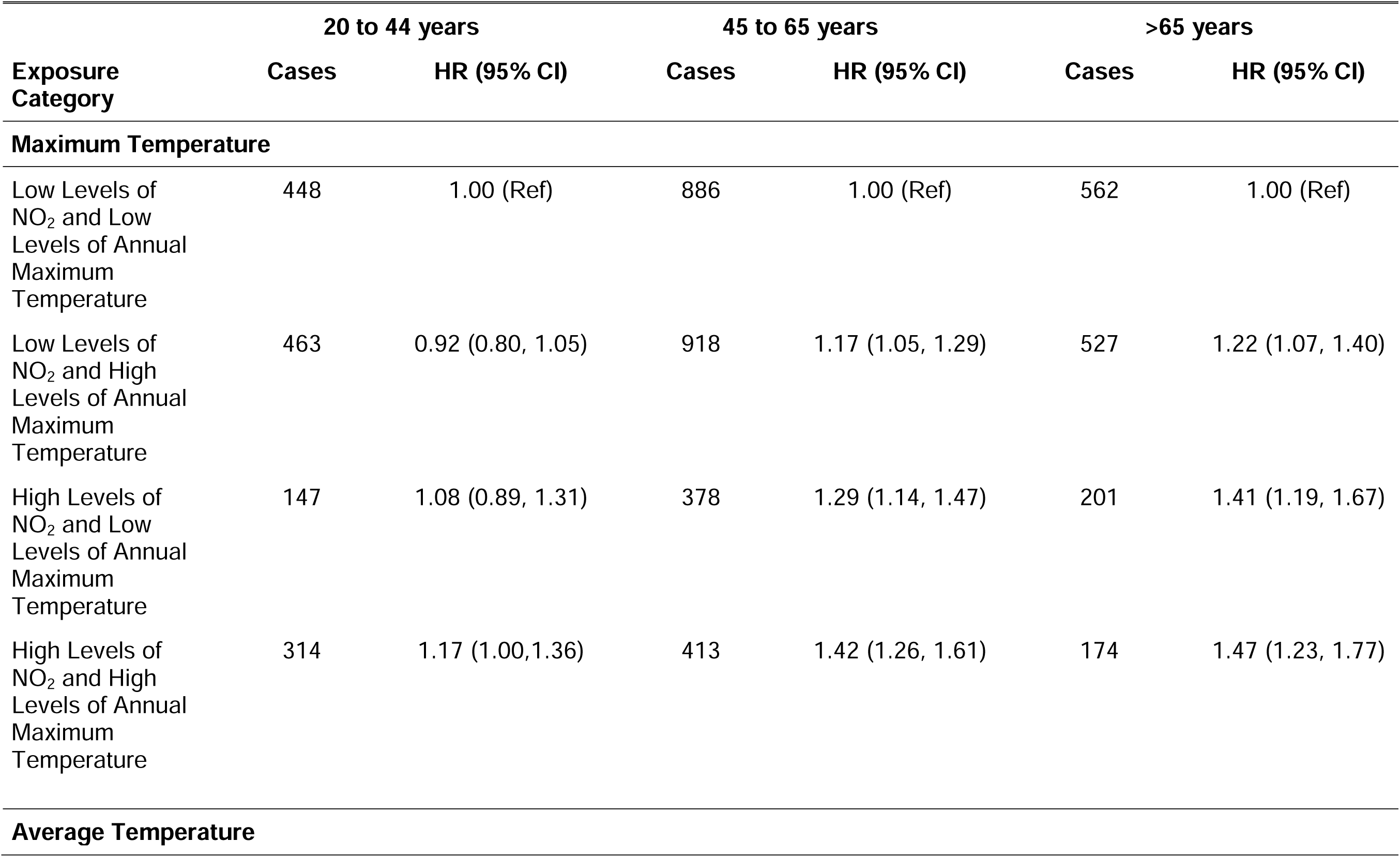
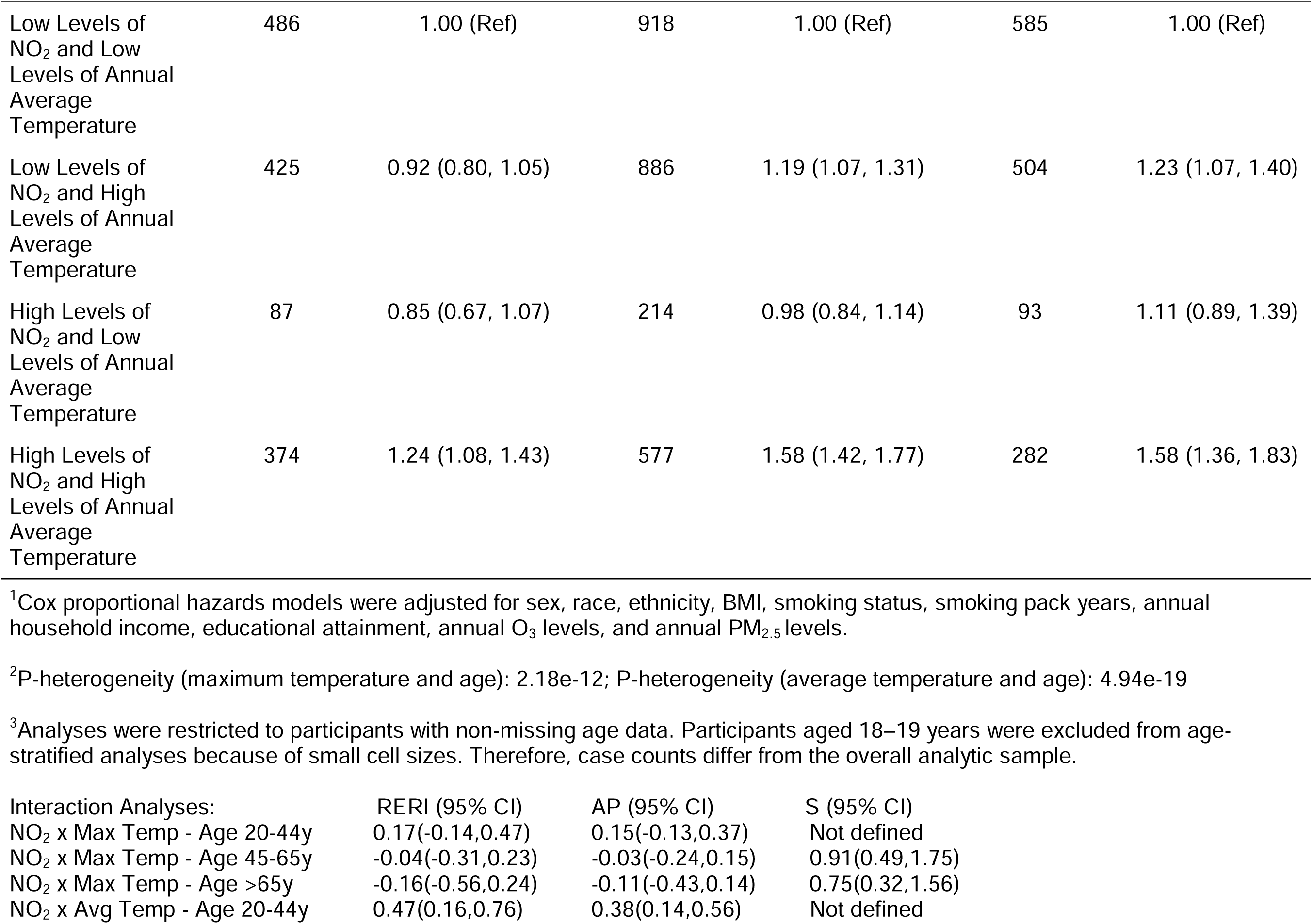
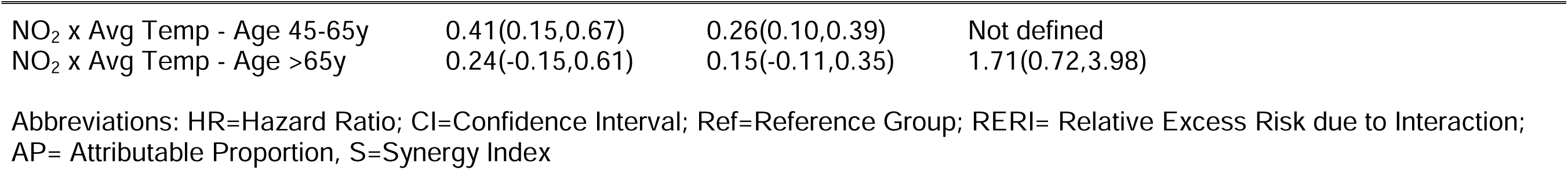
Associations of nitrogen dioxide and temperature with adult-onset asthma among various age groups in the All of Us Research Program. ^1, 2, 3^

|  | 20 to 44 years |  | 45 to 65 years |  | >65 years |  |
| --- | --- | --- | --- | --- | --- | --- |
| Exposure Category | Cases | HR (95% CI) | Cases | HR (95% CI) | Cases | HR (95% CI) |
| Maximum Temperature |  |  |  |  |  |  |
| Low Levels of NO<sub>2</sub> and Low Levels of Annual Maximum Temperature | 448 | 1.00 (Ref) | 886 | 1.00 (Ref) | 562 | 1.00 (Ref) |
| Low Levels of NO<sub>2</sub> and High Levels of Annual Maximum Temperature | 463 | 0.92 (0.80, 1.05) | 918 | 1.17 (1.05, 1.29) | 527 | 1.22 (1.07, 1.40) |
| High Levels of NO<sub>2</sub> and Low Levels of Annual Maximum Temperature | 147 | 1.08 (0.89, 1.31) | 378 | 1.29 (1.14, 1.47) | 201 | 1.41 (1.19, 1.67) |
| High Levels of NO<sub>2</sub> and High Levels of Annual Maximum Temperature | 314 | 1.17 (1.00,1.36) | 413 | 1.42 (1.26, 1.61) | 174 | 1.47 (1.23, 1.77) |
| Average Temperature |  |  |  |  |  |  |
| Low Levels of NO<sub>2</sub> and Low Levels of Annual Average Temperature | 486 | 1.00 (Ref) | 918 | 1.00 (Ref) | 585 | 1.00 (Ref) |
| Low Levels of NO<sub>2</sub> and High Levels of Annual Average Temperature | 425 | 0.92 (0.80, 1.05) | 886 | 1.19 (1.07, 1.31) | 504 | 1.23 (1.07, 1.40) |
| High Levels of NO<sub>2</sub> and Low Levels of Annual Average Temperature | 87 | 0.85 (0.67, 1.07) | 214 | 0.98 (0.84, 1.14) | 93 | 1.11 (0.89, 1.39) |
| High Levels of NO<sub>2</sub> and High Levels of Annual Average Temperature | 374 | 1.24 (1.08, 1.43) | 577 | 1.58 (1.42, 1.77) | 282 | 1.58 (1.36, 1.83) |

<sup>1</sup>Cox proportional hazards models were adjusted for sex, race, ethnicity, BMI, smoking status, smoking pack years, annual household income, educational attainment, annual O<sub>3</sub> levels, and annual PM<sub>2.5</sub> levels.
<sup>2</sup>P-heterogeneity (maximum temperature and age): 2.18e-12; P-heterogeneity (average temperature and age): 4.94e-19
<sup>3</sup>Analyses were restricted to participants with non-missing age data. Participants aged 18–19 years were excluded from age-stratified analyses because of small cell sizes. Therefore, case counts differ from the overall analytic sample.

| Interaction Analyses: | RERI (95% CI) | AP (95% CI) | S (95% CI) |
| --- | --- | --- | --- |
| NO<sub>2</sub> x Max Temp - Age 20-44y | 0.17(-0.14,0.47) | 0.15(-0.13,0.37) | Not defined |
| NO<sub>2</sub> x Max Temp - Age 45-65y | -0.04(-0.31,0.23) | -0.03(-0.24,0.15) | 0.91(0.49,1.75) |
| NO<sub>2</sub> x Max Temp - Age >65y | -0.16(-0.56,0.24) | -0.11(-0.43,0.14) | 0.75(0.32,1.56) |
| NO<sub>2</sub> x Avg Temp - Age 20-44y | 0.47(0.16,0.76) | 0.38(0.14,0.56) | Not defined |

|  |  |  |  |
| --- | --- | --- | --- |
| NO<sub>2</sub> x Avg Temp - Age 45-65y | 0.41(0.15,0.67) | 0.26(0.10,0.39) | Not defined |
| NO<sub>2</sub> x Avg Temp - Age >65y | 0.24(-0.15,0.61) | 0.15(-0.11,0.35) | 1.71(0.72,3.98) |

Abbreviations: HR=Hazard Ratio; CI=Confidence Interval; Ref=Reference Group; RERI= Relative Excess Risk due to Interaction; AP= Attributable Proportion, S=Synergy Index

### Income

We found that the joint associations of high NO_2_ × high maximum temperature and AOA risk were more apparent among certain socioeconomic subgroups **(Supplementary Table 3)**. The magnitude of the joint associations was highest among participants in the low annual household income category (<$75,000). Similar trends were observed when examining high NO_2_ × high average temperature, where the AOA risk increased with decreasing income. Here, the super-additive interaction was strongest in the low income group (NO_2_ x Avg Temp – Low: RERI=0.60 (0.30,0.90); AP=0.32 (0.17,0.45), **Supplementary Table 3**).

### Weighting to Account for Potential Volunteer Selection Bias

The observed joint associations were robust even after applying different weighting techniques to correct for potential volunteer selection bias from non-probabilistic sampling **(Supplementary Table 4).** Across all weighting methods, the association remained statistically significant. When examining conditions of high NO_2_ and high maximum temperature, higher AOA risk was observed in both weighting methods [(HR_Weight1_=1.26, 95%CI:1.12-1.41; HR_Weight2_=1.42, 95%CI:1.25-1.62)] compared to the reference groups. Similar trends were observed when examining high NO_2_ and high average temperature [(HR _Weight_ =1.35, 95%CI:1.21-1.50; HR _Weight2_=1.51, 95%CI:1.34-1.70)].

### Sensitivity Analyses – QaSIS

In the QaSIS-based quantile-survival screening analysis, the indicator variables representing NO_2_ × maximum temperature as well as NO_2_ × average temperature categories ranked between 6-8^th^, out of 18 predictors **(Supplementary Tables 5 and 6)**. These findings indicate that the joint association of NO_2_ × temperature was among the most influential variables for AOA cases with the earliest time-to-diagnosis.

### Sensitivity Analyses – CQR

Censored quantile regression analyses showed directionally consistent associations with the Cox models **(Supplementary Tables 7 and 8).** We found that high NO_2_ × high maximum temperature was associated with a non-significant earlier time-to-diagnosis (βτ = −0.22 (95% CI: −1.20, 0.66)); similar associations were found for high NO_2_ × high average temperature (βτ = −0.30 (95% CI: −1.27, 0.36)). These results do not show clear evidence that higher NO_2_ combined with higher temperature influences the most susceptible AOA cases differently than the overall incident cases.

## Discussion

In this large prospective cohort study of U.S. adults, we found that joint exposure to high NO_2_ and elevated temperature was associated with increased risk of AOA, with the additive interaction differing by temperature metric. Annual average temperature showed super-additive joint associations with NO_2_ overall, and this finding remained consistent across smoking strata, sex, age groups, and income levels. In contrast, annual maximum temperature did not show meaningful additive interaction with NO_2_ overall and had inconsistent patterns across subgroups, in spite of a strong Spearman correlation (r_s_=0.97) between maximum and average temperature. The consistency observed for sustained average temperature, but not for maximum temperature, suggests that cumulative heat exposure, rather than shorter periods of peak heat, is potentially more etiologically relevant for respiratory effects related to air pollution.

These findings are consistent with cumulative inflammatory and oxidative stress pathways rather than acute heat-stress responses. The super-additive interactions were most pronounced among ever smokers, lower-income participants, and younger adults, pointing toward differential susceptibility potentially from different exposure profiles, lifestyle exposures, and socioeconomic status. The robustness of these joint associations to adjustment for co-pollutants, atmospheric conditions, and forms of selection bias supports their interpretation as substantive findings.

While AOA risk was elevated in never smokers, the risk was even higher among individuals with a history of smoking. This finding is consistent with the understanding that smoking may increase the lungs’ vulnerability to environmental stressors, which amplifies inflammatory responses and impairs repair mechanisms (38, 39). It is also likely that the cumulative burden of smoking and air pollution leads to accelerated loss of lung function, causing the airways to be more susceptible to AOA triggers in adulthood (8, 40). Furthermore, the observed joint association of NO_2_ × temperature among never-smokers suggests that confounding by smoking was minimal. These findings build on recent research on non-smoking risk factors of AOA and contribute new insights into the risks associated with NO_2_ and temperature (41, 42).

We also observed heterogeneity in the joint associations of NO_2_ × temperature by sex, with a stronger association among males than females. However, the additive interaction of high NO_2_ and high average temperature was similar in magnitude for males and females. While asthma is generally more prevalent among adult females, previous research has shown that males may exhibit greater susceptibility to certain environmental exposures, potentially due to their differences in occupational exposures and lifestyle factors (43–45). These results highlight the need for comprehensive understanding of the disease across sexes.

We posit that the pathobiological mechanisms underlying the joint association between temperature and NO_2_ were primarily related to how heat exposure may enhance airway inflammation, increase respiratory absorption of pollutants, and alter immune responses to traffic-related pollutants such as NO_2_(46, 47). Our findings support temperature as an important co-exposure in the development of adult respiratory disease. In particular, long-term exposure to NO_2_ and heat, two common features of modern urban environments, may play a significant role in AOA pathogenesis (48, 49). As meteorological changes cause more frequent temperature extremes and potentially change pollution levels, understanding these exposures jointly becomes critical for informing targeted exposure reduction efforts (50).

This study has notable strengths. First, the AoU research program is a data-rich cohort of U.S. adults with linked EHRs and questionnaire data. The targeted recruitment of minority populations from across the United States allowed for detailed subgroup analyses with greater granularity and statistical power. Second, our use of satellite-based environmental data allowed for improved spatial resolution and consistency in estimating long-term NO₂, other chemical compounds, and temperature levels, enhancing the reliability of our findings.

Our study had some limitations. First, although we adjusted for many covariates, we cannot discount residual and unmeasured confounding. However, we applied rigorous confounder adjustment informed by prior literature and further evaluated the robustness of our findings through stratified and sensitivity analyses. Second, exposure assignment was based on residential three-digit ZIP codes to protect participant privacy, which may have masked the spatial heterogeneity of NO_2_ and led to exposure misclassification. However, such misclassification is likely non-differential with respect to AOA diagnoses and is expected to attenuate the observed associations toward the null. Additionally, due to data constraints, exposure estimates could only be assigned for a limited period of residential history. We recognize the potential for time-trend bias, as ambient NO_2_ concentrations generally decreased over the study period. Further, NO_2_ may not have been measured within the appropriate etiological window for AOA onset or diagnosis, since chronic respiratory diseases typically develop over long periods of cumulative exposure. Our NO_2_ estimates were based on residential addresses at enrollment, requiring the assumption that these exposures were representative of prior years. However, because the study population was largely residentially stable, with nearly two-thirds of the participants living at the current address for ≥3 years and almost 20% residing there for >20 years, we expect that our exposure estimates reflect etiologically meaningful periods for AOA development. To further minimize the potential for impacts on our associations, we applied long-term exposure estimates that incorporated multi-year annual averages, attempting to account for the cumulative burden of NO_2_ relevant to AOA pathogenesis.

In conclusion, we found that joint exposure to high NO_2_ and elevated temperature was associated with increased AOA risk, with super-additive joint associations observed consistently for annual average temperature, but not annual maximum temperature. Our findings suggest that sustained, rather than acute peak, heat exposure may be more etiologically relevant when considering joint air pollution and temperature effects on respiratory health. The interaction was most pronounced among ever smokers, lower-income participants, and younger adults. By integrating multiple environmental exposures, our study contributes to a more nuanced understanding of the air exposome and AOA risk. These findings have important implications for public health surveillance and targeted interventions, supporting the need to consider air pollution and cumulative heat exposure jointly rather than separately. However, given the limitations of our study, caution is warranted in interpreting these findings. External replication in other well-powered U.S. cohorts is needed to confirm our results.

## Conflict of Interest Statement

We declare no conflict(s) of interest.

## Data Availability

Controlled-access AoU population data are available at: https://www.researchallofus.org/. Air monitoring data are available at: https://search.earthdata.nasa.gov/. Atmospheric temperature data are found here: https://www.ncei.noaa.gov/access.

https://www.ncei.noaa.gov/access

https://search.earthdata.nasa.gov

## Acknowledgements

This work was funded by the intramural research program of the National Institutes of Health – National Heart, Lung, and Blood Institute. This work used computational resources of the NIH STRIDES Initiative (https://cloud.nih.gov) through the other transaction agreement (GCP: OT2OD027060, Azure: OT2OD032100).The contributions of the NIH author(s) are considered Works of the United States Government. The findings and conclusions presented in this paper are those of the author(s) and do not necessarily reflect the views of the NIH or the U.S. Department of Health and Human Services.

We extend our appreciation to Dr. Véronique Roger for her support. Lastly, we thank all the participants in the All of Us research program.

All authors have seen and approved the manuscript.

## Ethics Statement

The All of Us Research Program was reviewed by the Institutional Review Board (IRB) of the All of Us Research Program, and approved by the IRB at the National Institutes of Health. All participants provided informed consent. The All of Us IRB follows the regulations and guidance of the NIH Office for Human Research Protections for all studies, ensuring that the rights and welfare of research participants are overseen and protected uniformly.

## Author Contributions

S.L. led the study, conducted the primary analyses, and wrote the initial manuscript draft. G.A.G. provided bioinformatic and programming support. H.W. conducted the American Community Survey weighted analyses. J.L. served as the study manager. S.V.C. provided programming support. J.Y.Y.W. supervised the study. J.A.F., M.H., and R.R.J. contributed to manuscript review and editing. All authors reviewed and edited the manuscript.

## Generative Artificial Intelligence Declaration

Generative AI tools (Claude, Anthropic; ChatGPT, OpenAI) were used solely for editing and error detection. The authors take full responsibility for all content.

## Supplementary Tables and Figures

**Supplementary Table 1:** Overall associations between NO_2_ and adult-onset asthma risk in the All of Us Research Program. ^1, 2^

|  |  |  |
| --- | --- | --- |
| **Nitrogen dioxide (NO<sub>2</sub>) Concentrations** | **Cases** | **HR (95% CI)** |
| Q1 (4.14 – ≤18.31 ppb) | 2150 | 1.00 (Ref) |
| Q2 (>18.31 – ≤27.20 ppb) | 2687 | 0.98 (0.92, 1.03) |
| Q3 (>27.20 – ≤31.21 ppb) | 1936 | 0.85 (0.80, 0.91) |
| Q4 (>31.21 – 44.69 ppb) | 2859 | 1.08 (1.02, 1.15) |
| Continuous (per 8 ppb) | 9632 | 1.03 (1.01, 1.04)<br>P-trend=0.006 |
| <sup>1</sup>Cox proportional hazards models were adjusted for age, sex, race, ethnicity, BMI, smoking status, smoking pack years, annual household income, educational attainment, annual O<sub>3</sub> levels, and annual PM<sub>2.5</sub> levels.<br><sup>2</sup>Analyses involving NO<sub>2</sub> quartiles and continuous exposure models were restricted to participants with complete data.<br>Abbreviations: HR=Hazard Ratio; CI=Confidence Interval; Ref=Reference Group |  |  |

**Supplementary Table 2:** Overall associations between temperature and adult-onset asthma risk in the All of Us Research Program. ^1, 2^

| Exposure Category | Cases | HR (95% CI) |
| --- | --- | --- |
| Annual Maximum Temperature (per 1°C) | 9632 | 1.01 (1.00, 1.02)<br>P-trend=0.035 |
| Annual Average Temperature (per 1°C) | 9632 | 1.01 (1.00, 1.01)<br>P-trend=0.074 |
| <sup>1</sup>Cox proportional hazards models were adjusted for age, sex, race, ethnicity, BMI, smoking status, smoking pack years, annual household income, educational attainment, annual O<sub>3</sub> levels, and annual PM<sub>2.5</sub> levels<br><sup>2</sup>Analyses involving quartiles and continuous exposure models were restricted to participants with complete data, resulting in a reduced analytic sample compared to the overall study population.<br>Abbreviations: HR=Hazard Ratio; CI=Confidence Interval |  |  |

**Supplementary Table 3:**
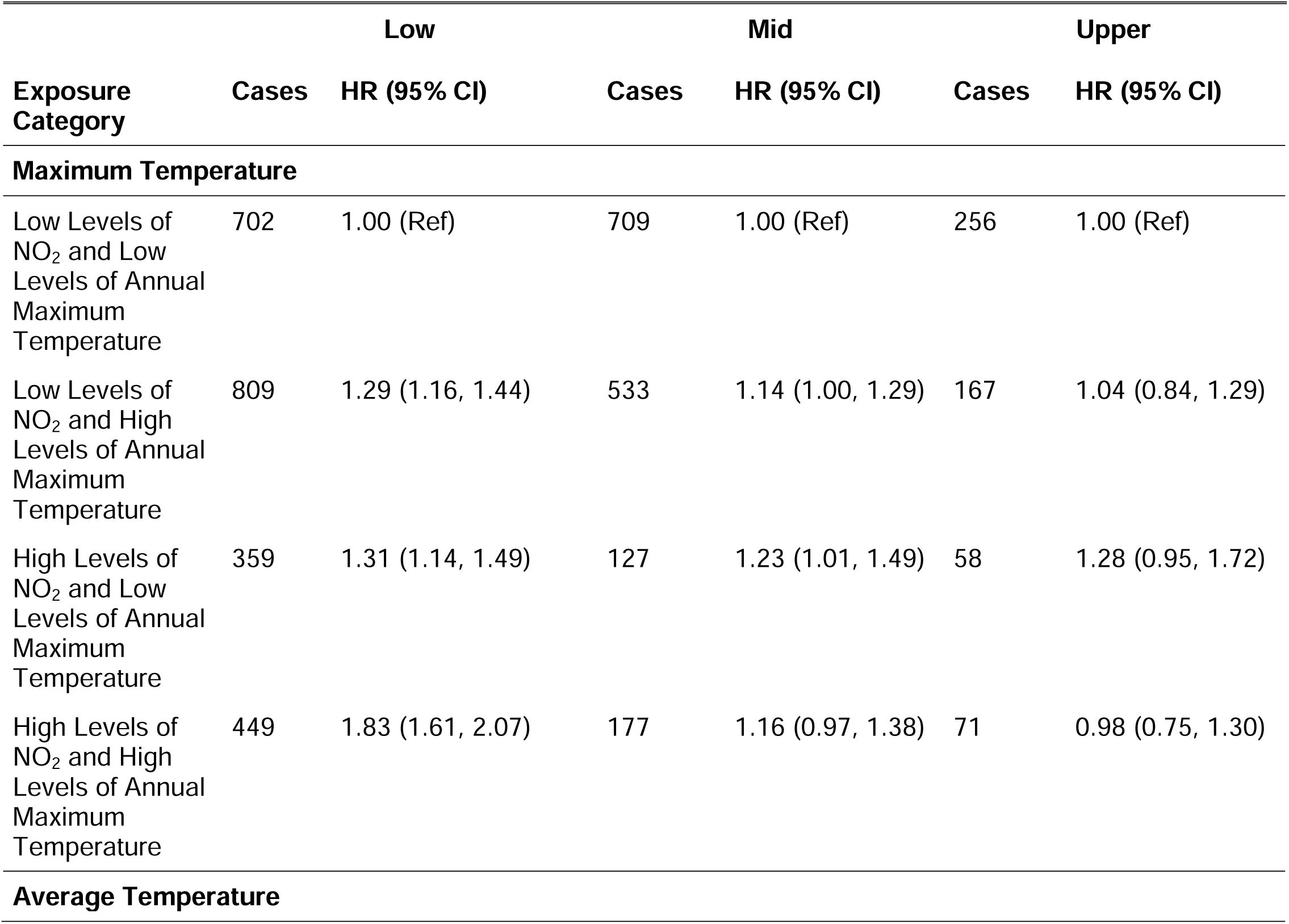
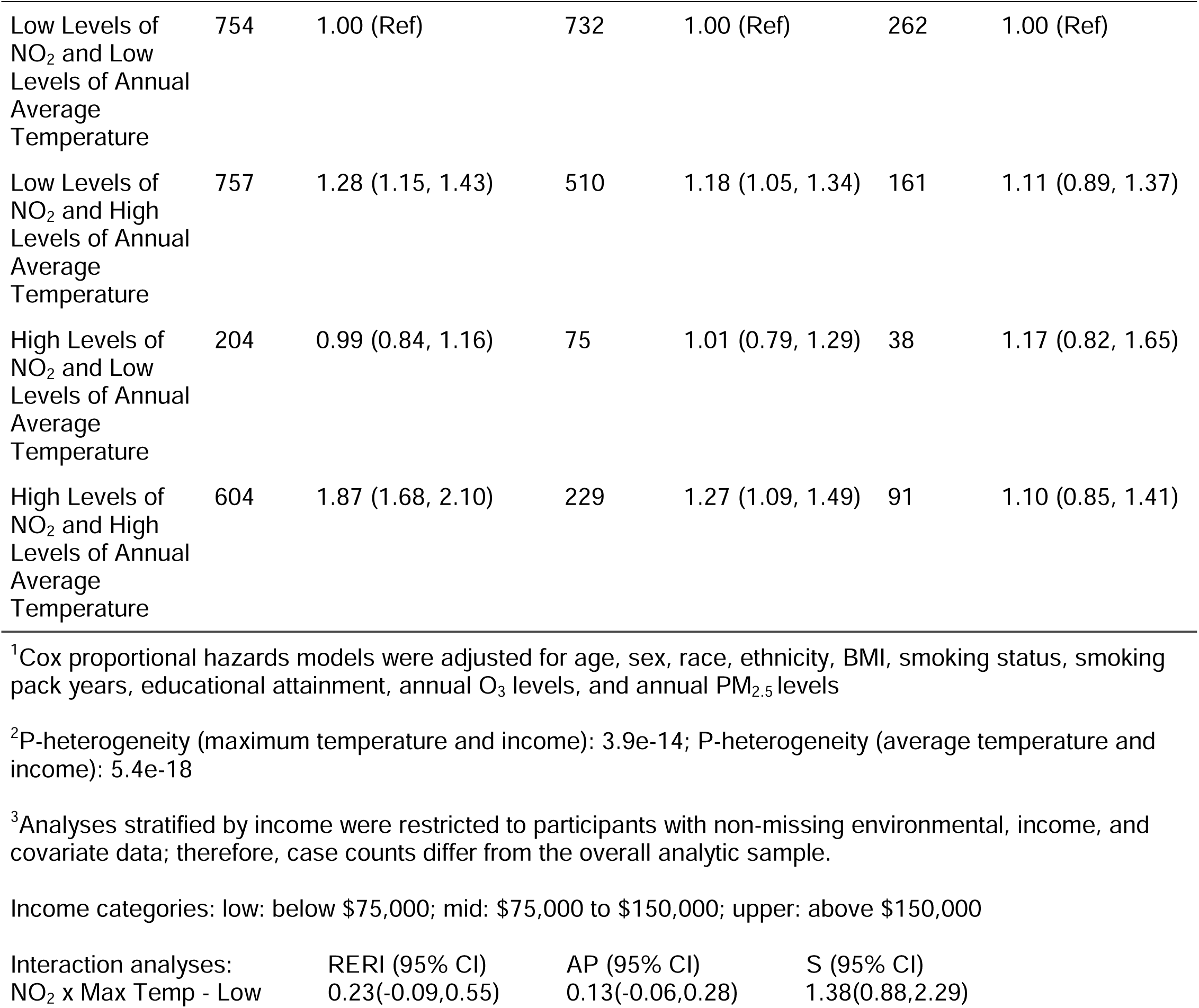
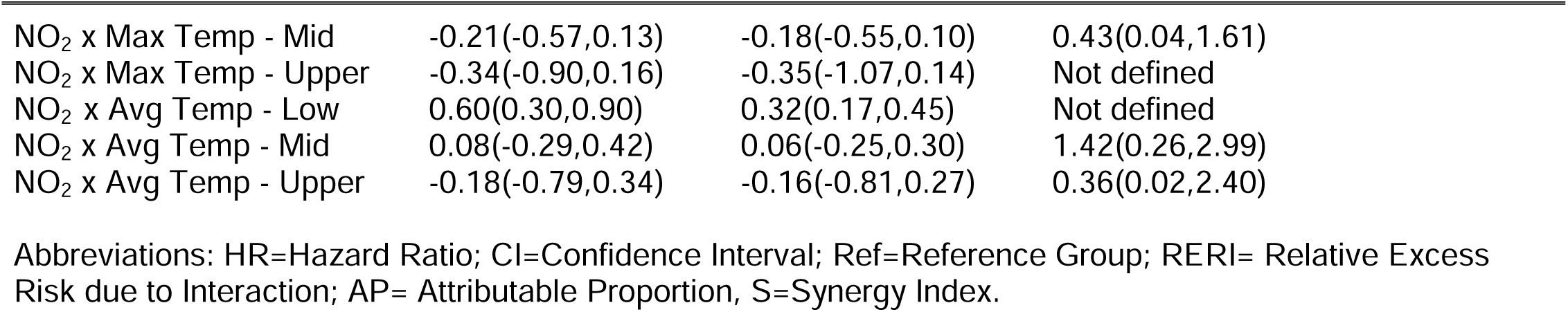
Associations of NO_2_ and temperature with adult-onset asthma among various average annual household income groups in the All of Us Research Program. ^1, 2, 3^

| Exposure Category | Low |  | Mid |  | Upper |  |
| --- | --- | --- | --- | --- | --- | --- |
|  | Cases | HR (95% CI) | Cases | HR (95% CI) | Cases | HR (95% CI) |
| Maximum Temperature |  |  |  |  |  |  |
| Low Levels of NO<sub>2</sub> and Low Levels of Annual Maximum Temperature | 702 | 1.00 (Ref) | 709 | 1.00 (Ref) | 256 | 1.00 (Ref) |
| Low Levels of NO<sub>2</sub> and High Levels of Annual Maximum Temperature | 809 | 1.29 (1.16, 1.44) | 533 | 1.14 (1.00, 1.29) | 167 | 1.04 (0.84, 1.29) |
| High Levels of NO<sub>2</sub> and Low Levels of Annual Maximum Temperature | 359 | 1.31 (1.14, 1.49) | 127 | 1.23 (1.01, 1.49) | 58 | 1.28 (0.95, 1.72) |
| High Levels of NO<sub>2</sub> and High Levels of Annual Maximum Temperature | 449 | 1.83 (1.61, 2.07) | 177 | 1.16 (0.97, 1.38) | 71 | 0.98 (0.75, 1.30) |
| Average Temperature |  |  |  |  |  |  |
| Low Levels of NO<sub>2</sub> and Low Levels of Annual Average Temperature | 754 | 1.00 (Ref) | 732 | 1.00 (Ref) | 262 | 1.00 (Ref) |
| Low Levels of NO<sub>2</sub> and High Levels of Annual Average Temperature | 757 | 1.28 (1.15, 1.43) | 510 | 1.18 (1.05, 1.34) | 161 | 1.11 (0.89, 1.37) |
| High Levels of NO<sub>2</sub> and Low Levels of Annual Average Temperature | 204 | 0.99 (0.84, 1.16) | 75 | 1.01 (0.79, 1.29) | 38 | 1.17 (0.82, 1.65) |
| High Levels of NO<sub>2</sub> and High Levels of Annual Average Temperature | 604 | 1.87 (1.68, 2.10) | 229 | 1.27 (1.09, 1.49) | 91 | 1.10 (0.85, 1.41) |

<sup>1</sup>Cox proportional hazards models were adjusted for age, sex, race, ethnicity, BMI, smoking status, smoking pack years, educational attainment, annual O<sub>3</sub> levels, and annual PM<sub>2.5</sub> levels
<sup>2</sup>P-heterogeneity (maximum temperature and income): 3.9e-14; P-heterogeneity (average temperature and income): 5.4e-18
<sup>3</sup>Analyses stratified by income were restricted to participants with non-missing environmental, income, and covariate data; therefore, case counts differ from the overall analytic sample.

Income categories: low: below \$75,000; mid: \$75,000 to \$150,000; upper: above \$150,000
|  |  |  |  |
| --- | --- | --- | --- |
| Interaction analyses: | RERI (95% CI) | AP (95% CI) | S (95% CI) |
| NO<sub>2</sub> x Max Temp - Low | 0.23(-0.09,0.55) | 0.13(-0.06,0.28) | 1.38(0.88,2.29) |

|  |  |  |  |
| --- | --- | --- | --- |
| NO<sub>2</sub> x Max Temp - Mid | -0.21(-0.57,0.13) | -0.18(-0.55,0.10) | 0.43(0.04,1.61) |
| NO<sub>2</sub> x Max Temp - Upper | -0.34(-0.90,0.16) | -0.35(-1.07,0.14) | Not defined |
| NO<sub>2</sub> x Avg Temp - Low | 0.60(0.30,0.90) | 0.32(0.17,0.45) | Not defined |
| NO<sub>2</sub> x Avg Temp - Mid | 0.08(-0.29,0.42) | 0.06(-0.25,0.30) | 1.42(0.26,2.99) |
| NO<sub>2</sub> x Avg Temp - Upper | -0.18(-0.79,0.34) | -0.16(-0.81,0.27) | 0.36(0.02,2.40) |

Abbreviations: HR=Hazard Ratio; CI=Confidence Interval; Ref=Reference Group; RERI= Relative Excess Risk due to Interaction; AP= Attributable Proportion, S=Synergy Index.

**Supplementary Table 4:**
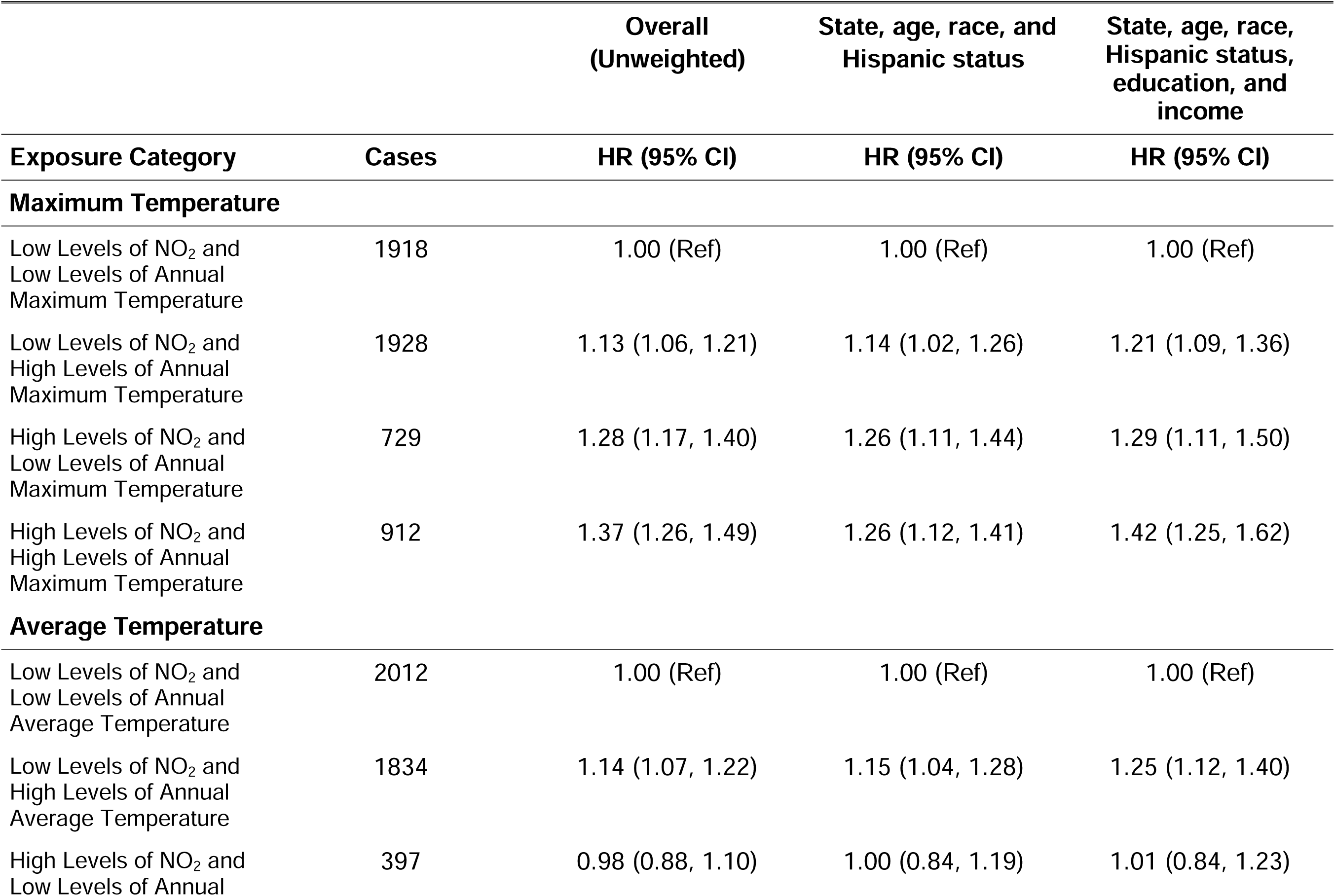
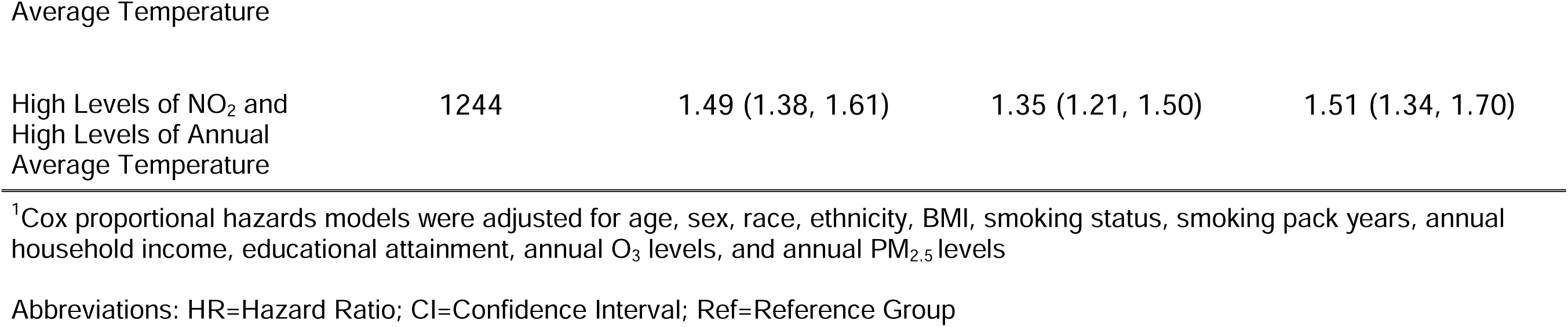
Nitrogen dioxide, temperature, and adult-onset asthma risk in the All of Us Research Program: application of cell weighting to account for non-probabilistic sampling. ^1^

|  |  | Overall<br>(Unweighted) | State, age, race, and<br>Hispanic status | State, age, race,<br>Hispanic status,<br>education, and<br>income |
| --- | --- | --- | --- | --- |
| Exposure Category | Cases | HR (95% CI) | HR (95% CI) | HR (95% CI) |
| **Maximum Temperature** |  |  |  |  |
| Low Levels of NO<sub>2</sub> and<br>Low Levels of Annual<br>Maximum Temperature | 1918 | 1.00 (Ref) | 1.00 (Ref) | 1.00 (Ref) |
| Low Levels of NO<sub>2</sub> and<br>High Levels of Annual<br>Maximum Temperature | 1928 | 1.13 (1.06, 1.21) | 1.14 (1.02, 1.26) | 1.21 (1.09, 1.36) |
| High Levels of NO<sub>2</sub> and<br>Low Levels of Annual<br>Maximum Temperature | 729 | 1.28 (1.17, 1.40) | 1.26 (1.11, 1.44) | 1.29 (1.11, 1.50) |
| High Levels of NO<sub>2</sub> and<br>High Levels of Annual<br>Maximum Temperature | 912 | 1.37 (1.26, 1.49) | 1.26 (1.12, 1.41) | 1.42 (1.25, 1.62) |
| **Average Temperature** |  |  |  |  |
| Low Levels of NO<sub>2</sub> and<br>Low Levels of Annual<br>Average Temperature | 2012 | 1.00 (Ref) | 1.00 (Ref) | 1.00 (Ref) |
| Low Levels of NO<sub>2</sub> and<br>High Levels of Annual<br>Average Temperature | 1834 | 1.14 (1.07, 1.22) | 1.15 (1.04, 1.28) | 1.25 (1.12, 1.40) |
| High Levels of NO<sub>2</sub> and<br>Low Levels of Annual | 397 | 0.98 (0.88, 1.10) | 1.00 (0.84, 1.19) | 1.01 (0.84, 1.23) |
| Average Temperature |  |  |  |  |
| High Levels of NO<sub>2</sub> and<br>High Levels of Annual<br>Average Temperature | 1244 | 1.49 (1.38, 1.61) | 1.35 (1.21, 1.50) | 1.51 (1.34, 1.70) |

<sup>1</sup>Cox proportional hazards models were adjusted for age, sex, race, ethnicity, BMI, smoking status, smoking pack years, annual household income, educational attainment, annual O<sub>3</sub> levels, and annual PM<sub>2.5</sub> levels
Abbreviations: HR=Hazard Ratio; CI=Confidence Interval; Ref=Reference Group

**Supplementary Table 5.**
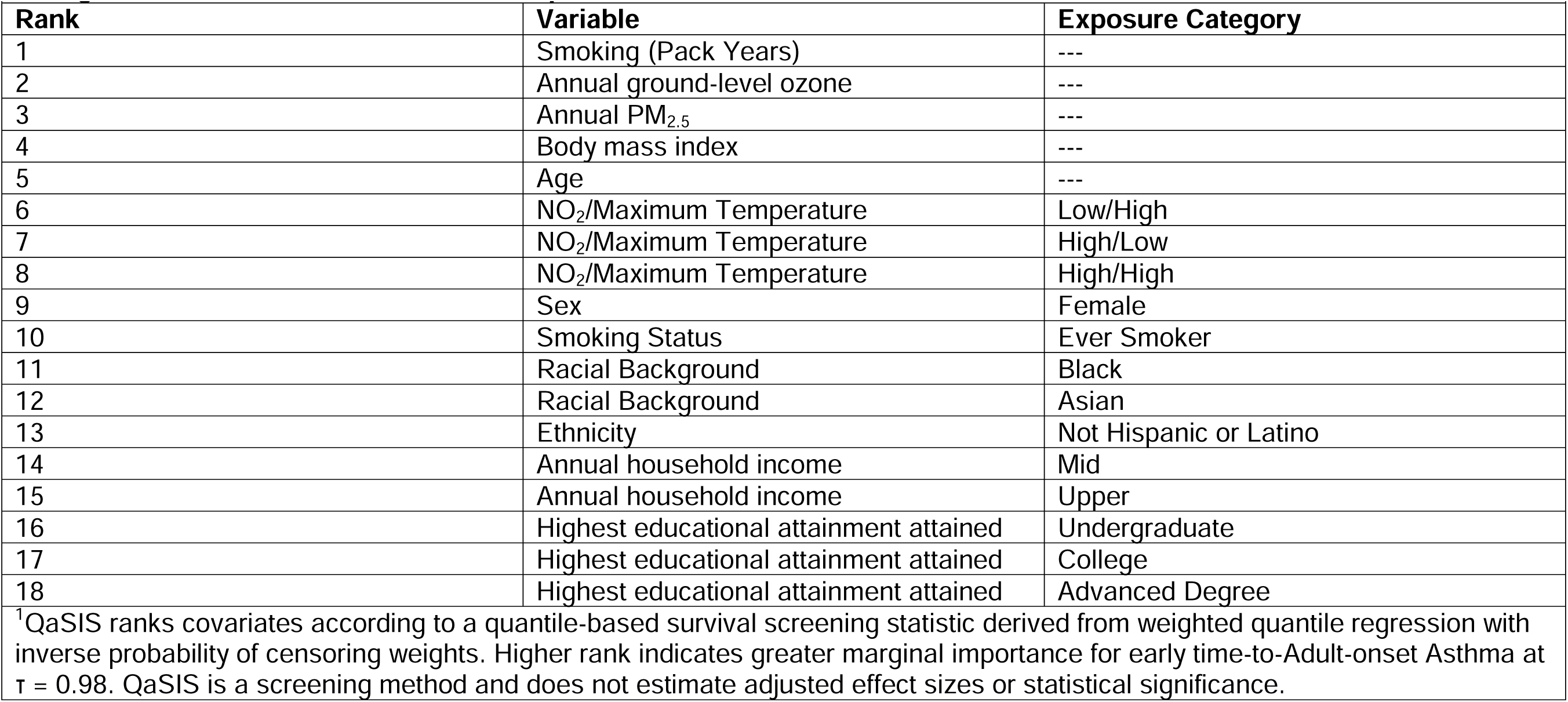
Quantile-Adaptive Survival Screening (QaSIS) Results for Adult-onset Asthma Evaluating Nitrogen Dioxide (NO_2_) and Maximum Temperature^1^.

**Supplementary Table 6.** Quantile-Adaptive Survival Screening (QaSIS) Results for Adult-onset Asthma Evaluating NO_2_ and Average Temperature^1^.

| Rank | Variable | Exposure Category |
| --- | --- | --- |
| 1 | Smoking (Pack Years) | --- |
| 2 | Annual ground-level ozone | --- |
| 3 | Annual PM<sub>2.5</sub> | --- |
| 4 | Body mass index | --- |
| 5 | Age | --- |
| 6 | NO<sub>2</sub>/Average Temperature | Low/High |
| 7 | NO<sub>2</sub>/Average Temperature | High/Low |
| 8 | NO<sub>2</sub>/Average Temperature | High/High |
| 9 | Sex | Female |
| 10 | Smoking Status | Ever Smoker |
| 11 | Racial Background | Black |
| 12 | Racial Background | Asian |
| 13 | Ethnicity | Not Hispanic or Latino |
| 14 | Annual household income | Mid |
| 15 | Annual household income | Upper |
| 16 | Highest educational attainment attained | Undergraduate |
| 17 | Highest educational attainment attained | College |
| 18 | Highest educational attainment attained | Advanced Degree |

<sup>1</sup>QaSIS ranks covariates according to a quantile-based survival screening statistic derived from weighted quantile regression with inverse probability of censoring weights. Higher rank indicates greater marginal importance for early time-to-Adult-onset Asthma at $\tau = 0.98$ .

**Supplementary Table 7:** Nitrogen dioxide (NO_2_), maximum temperature, and adult-onset asthma risk in the All of Us Research Program: censored quantile regression method ^1, 2^.

| Exposure Category | $\beta\tau$ (95% CI) |
| --- | --- |
| Low Levels of NO<sub>2</sub> and Low Levels of Annual Maximum Temperature | 0.00 (Ref) |
| Low Levels of NO<sub>2</sub> and High Levels of Annual Maximum Temperature | -0.12 (-0.52, 0.83) |
| High Levels of NO<sub>2</sub> and Low Levels of Annual Maximum Temperature | -0.26 (-1.51, 0.40) |
| High Levels of NO<sub>2</sub> and High Levels of Annual Maximum Temperature | -0.22 (-1.20, 0.66) |
| <sup>1</sup>CQR models were adjusted for age, sex, race, ethnicity, BMI, smoking status, smoking pack years, annual household income, educational attainment, annual O<sub>3</sub> levels, and annual PM<sub>2.5</sub> levels<br>Abbreviations: CI=Confidence Interval; $\beta\tau$=Beta Tau; Ref=Reference Group<br><sup>2</sup>Maximum Temperature (tau = 0.02) | |

**Supplementary Table 8:** NO2, average temperature, and adult-onset asthma risk in the All of Us Research Program utilizing a censored quantile regression method^1, 2^.

| Exposure Category | $\beta_{\tau}$ (95% CI) |
| --- | --- |
| Low Levels of NO<sub>2</sub> and Low Levels of Annual Average Temperature | 0.00 (Ref) |
| Low Levels of NO<sub>2</sub> and High Levels of Annual Average Temperature | -0.14 (-0.78, 0.55) |
| High Levels of NO<sub>2</sub> and Low Levels of Annual Average Temperature | 0.03 (-1.70, 0.59) |
| High Levels of NO<sub>2</sub> and High Levels of Annual Average Temperature | -0.30 (-1.27, 0.36) |
| <sup>1</sup>CQR methods were adjusted for age, sex, race, ethnicity, BMI, smoking status, smoking pack years, annual household income, educational attainment, annual O<sub>3</sub> levels, and annual PM<sub>2.5</sub> levels.<br>Abbreviations: CI=Confidence Interval; $\beta_{\tau}$=Beta Tau; Ref=Reference Group<br><sup>2</sup>Average Temperature (tau = 0.02) |  |

**Supplementary Figure 1:**
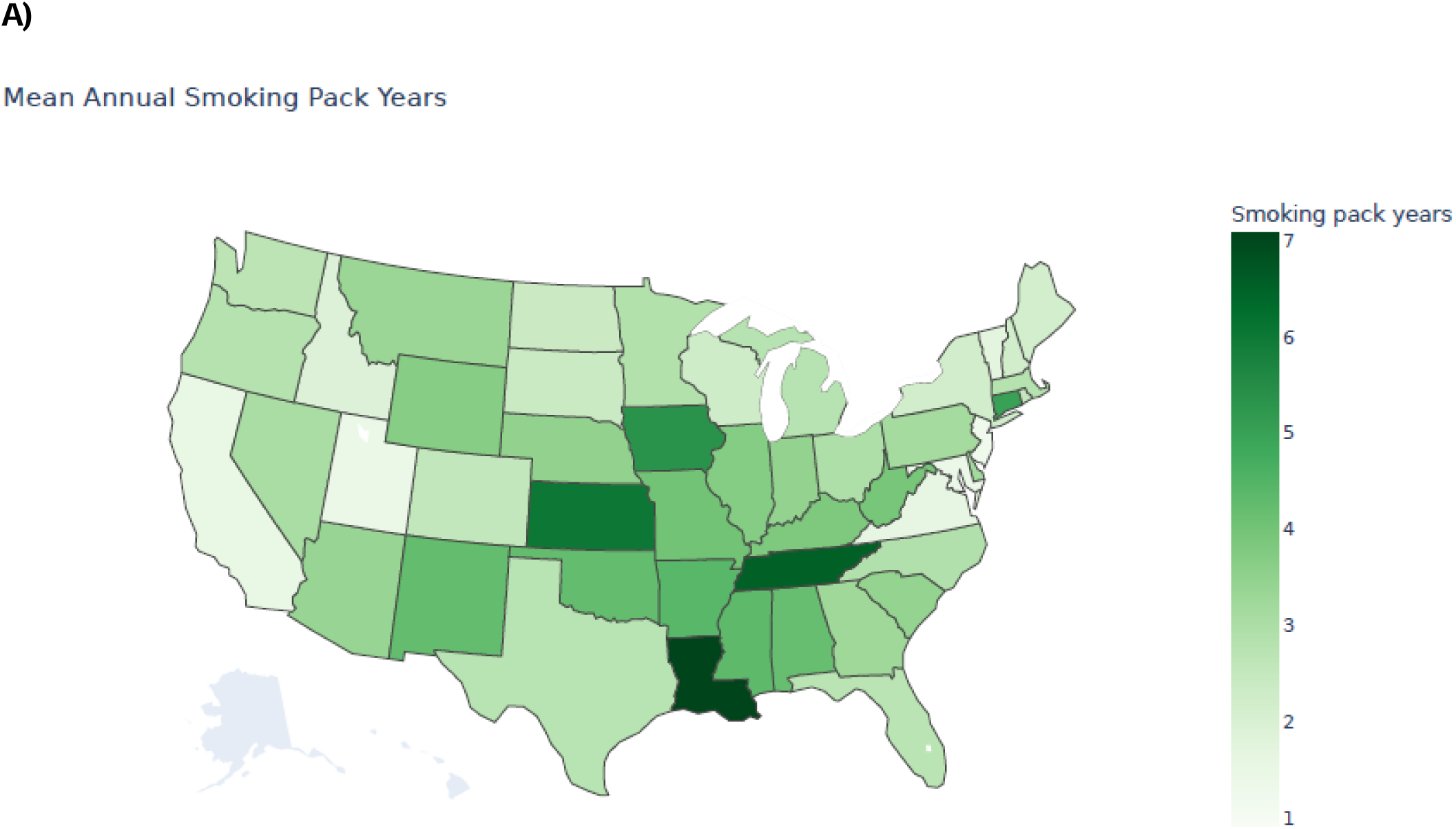
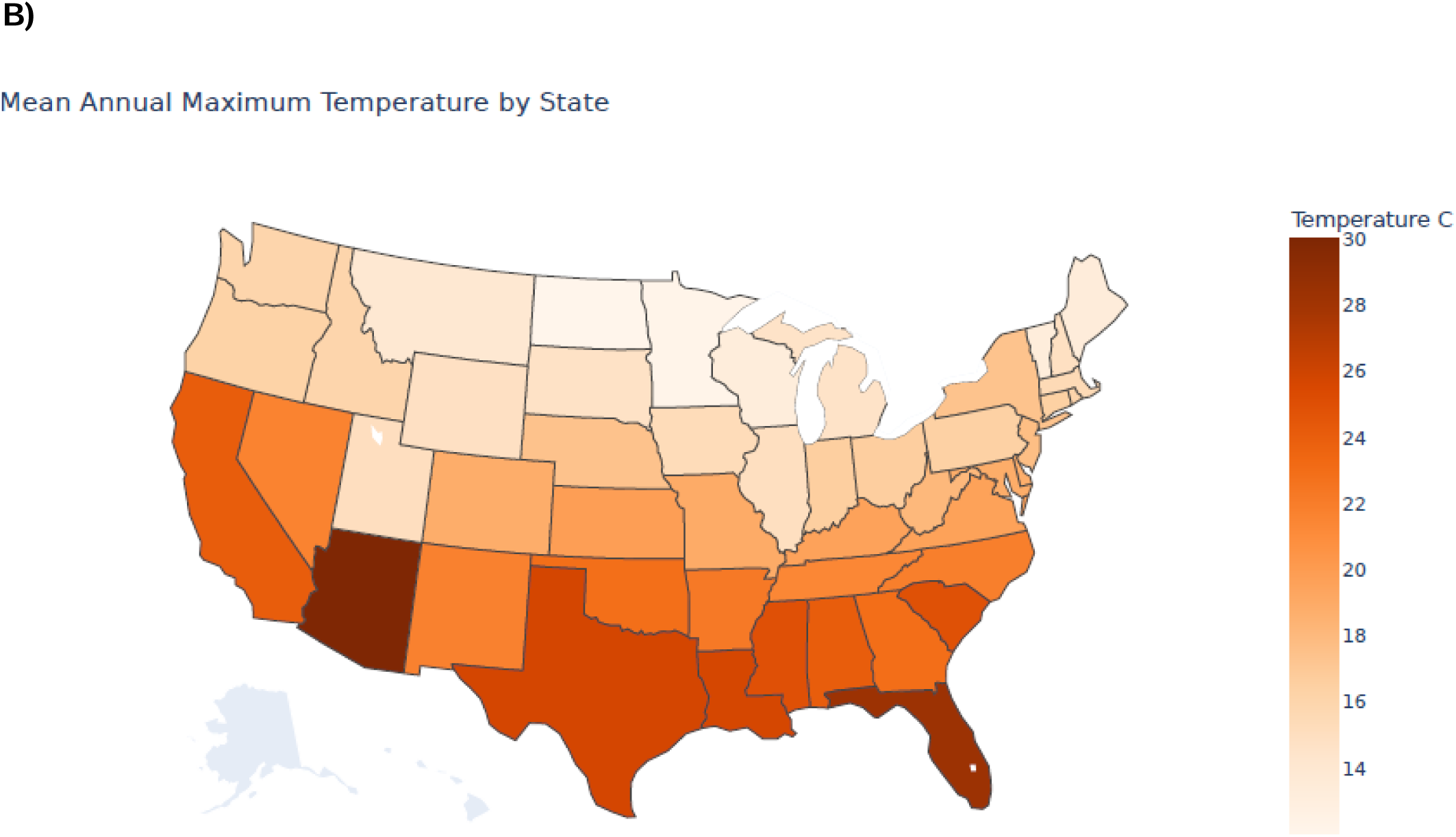
Distribution of A) annual smoking pack years and B) annual maximum temperature by state in the contiguous United States

## Notes

### Competing Interest Statement

The authors have declared no competing interest.

